# Persistent symptoms following SARS-CoV-2 infection in a random community sample of 508,707 people

**DOI:** 10.1101/2021.06.28.21259452

**Authors:** Matthew Whitaker, Joshua Elliott, Marc Chadeau-Hyam, Steven Riley, Ara Darzi, Graham Cooke, Helen Ward, Paul Elliott

## Abstract

**Background:** Long COVID, describing the long-term sequelae after SARS-CoV-2 infection, remains a poorly defined syndrome. There is uncertainty about its predisposing factors and the extent of the resultant public health burden, with estimates of prevalence and duration varying widely.

**Methods:** Within rounds 3–5 of the REACT-2 study, 508,707 people in the community in England were asked about a prior history of COVID-19 and the presence and duration of 29 different symptoms. We used uni-and multivariable models to identify predictors of persistence of symptoms (12 weeks or more). We estimated the prevalence of symptom persistence at 12 weeks, and used unsupervised learning to cluster individuals by symptoms experienced.

**Findings:** Among the 508,707 participants, the weighted prevalence of self-reported COVID-19 was 19.2% (95% CI: 19.1,19.3). 37.7% of 76,155 symptomatic people post COVID-19 experienced at least one symptom, while 14.8% experienced three or more symptoms, lasting 12 weeks or more. This gives a weighted population prevalence of persistent symptoms of 5.75% (5.68, 5.81) for one and 2.22% (2.1, 2.26) for three or more symptoms. Almost a third of people (8,771/28,713 [30.5%]) with at least one symptom lasting 12 weeks or more reported having had severe COVID-19 symptoms (“significant effect on my daily life”) at the time of their illness, giving a weighted prevalence overall for this group of 1.72% (1.69,1.76). The prevalence of persistent symptoms was higher in women than men (OR: 1.51 [1.46,1.55]) and, conditional on reporting symptoms, risk of persistent symptoms increased linearly with age by 3.5 percentage points per decade of life. Obesity, smoking or vaping, hospitalisation, and deprivation were also associated with a higher probability of persistent symptoms, while Asian ethnicity was associated with a lower probability. Two stable clusters were identified based on symptoms that persisted for 12 weeks or more: in the largest cluster, tiredness predominated, while in the second there was a high prevalence of respiratory and related symptoms.

**Interpretation:** A substantial proportion of people with symptomatic COVID-19 go on to have persistent symptoms for 12 weeks or more, which is age-dependent. Clinicians need to be aware of the differing manifestations of Long COVID which may require tailored therapeutic approaches. Managing the long-term sequelae of SARS-CoV-2 infection in the population will remain a major challenge for health services in the next stage of the pandemic.

**Funding:** The study was funded by the Department of Health and Social Care in England.

**Research in context:** *Evidence before this study:* Recent systematic reviews have documented the wide range of symptoms and reported prevalence of persistent symptoms following COVID-19. A dynamic review of Long COVID studies (NIHR Evidence) in March 2021 summarised the literature on the prevalence of persistent symptoms after acute COVID19, and reported that most studies (14) were of hospitalised patients, with higher prevalence of persistent symptoms compared with two community-based studies. There was limited evidence from community studies beyond 12 weeks. Another systematic review reported a median of over 70% of people with symptoms lasting at least 60 days. A review of risk factors for Long COVID found consistent evidence for an increased risk amongst women and those with high body mass index (BMI) but inconsistent findings on the role of age and little evidence concerning risks among different socioeconomic or ethnic groups which are often not well captured in routine healthcare records. Long COVID is increasingly recognised as heterogenous, likely underpinned by differing biological mechanisms, but there is not yet consensus on defining subtypes of the condition.

*Added value of this study:* This community-based study of over half a million people was designed to be representative of the adult population of England. A random sample of adults ages 18 years and above registered with a GP were invited irrespective of previous access to services for COVID-19, providing an estimate of population prevalence that was representative of the whole population. The findings show substantial declines in symptom prevalence over the first 12 weeks following Covid-19, reported by nearly one fifth of respondents, of whom over a third remained symptomatic at 12 weeks and beyond, with little evidence for decline thereafter. Risk factors identified for persistent symptoms (12 weeks or more) suggestive of Long COVID confirm some previous findings - an increased risk in women, obese and overweight individuals and those hospitalised for COVID-19, with strong evidence for an increasing risk with age. Additional evidence was found for an increased risk in those with lower income, smoking or vaping and healthcare or care home workers. A lower risk was found in those of Asian ethnicity. Clustering identified two distinct groups of individuals <u>wit</u>h different symptom profiles at 12 weeks, highlighting the heterogeneity of clinical presentation. The smaller cluster had higher prevalence of respiratory and related symptoms, while for those in the larger cluster tiredness was the dominant symptom, with lower prevalence of organ-specific symptoms.

*Implications of available evidence:* There is a high prevalence of persistent symptoms beyond 12 weeks after acute COVID-19, with little evidence of decline thereafter. This highlights the needs for greater support for patients, both through specialised services and, for those from low-income settings, financial support. The understanding that there are distinct clusters of persistent symptoms, the most common of which is dominated by fatigue, is important for the recognition and clinical management of the condition outside of specialised services.

## Introduction

The UK has experienced one of the largest epidemics of COVID-19 in Europe. As a new disease, the natural history beyond the immediate illness and the possible long-term sequelae remain largely unknown. As well as the acute risk of hospitalisation and death from COVID-19, some people who develop symptoms have a prolonged and debilitating illness that may continue for weeks or months (so-called Long COVID or post-COVID syndrome).^1–5^ Patients sharing their experience on social media and establishing support groups were key to raising awareness of persistent symptoms and coined the term Long COVID.

The frequency and duration of persistent symptoms from COVID-19 are poorly understood and represent a major knowledge gap if effective treatments and management strategies are to be developed. Reported symptoms/signs include severe fatigue, breathlessness, chest pain or heaviness, fever, palpitations, cognitive impairment (‘brain fog’), anosmia, ageusia, skin rash, joint pain or swelling. ^1–5^ Estimates of symptom prevalence and persistence vary substantially, arguably due to heterogeneous study designs and syndrome definitions.^6–9^ It has been suggested that Long COVID describes a group of disparate conditions, including post-viral syndromes, long-term tissue or organ damage and ongoing inflammation.^3,7,10,11^

Occurrence of Long COVID appears to be associated with the severity of acute COVID-19 symptoms; for example, high prevalence of persistent symptoms has been reported among people hospitalised with COVID-19.^12,13^ The number of acute symptoms has also been associated with risk of long COVID, alongside older age and female sex.^6^

While many Long COVID studies so far have focused on hospitalised COVID-19 cases,^12–16^ here we report data from random community-based samples of the population in England. These involved over half a million people who took part in rounds three to five of the Real-Time Assessment of Community Transmission-2 (REACT-2) study between September 2020 and February 2021. We estimate symptom prevalence and investigate co-occurrence of symptoms among participants reporting symptoms lasting 12 weeks or more following suspected or confirmed COVID-19.

## Methods

### Participants

The REACT-2 programme evaluates community prevalence of SARS-CoV-2 anti-spike protein antibody positivity in England. Random population samples of adults in England were invited to take part every 2–4 months using the National Health Service (NHS) patient list to achieve similar numbers of participants in each of 315 lower-tier local authority (LTLA) areas.^17^ Participants registered via an online portal or by telephone. Those registered were sent a test kit by post that included a self-administered point-of-care lateral flow immunoassay (LFIA) test with instructions and a link to an online video. Participants completed a survey (online/telephone) upon completion of their self-test. Participants provided information on demographics, household composition, whether or not they thought that they had had COVID-19, whether or not they had had a PCR test, comorbidities, symptoms related to COVID-19, severity of symptoms, and duration of any of a list of 29 symptoms.^18^ In addition, we asked participants to report any other symptoms in free text. Personalised invitations were sent to between 560,000 and 600,000 individuals aged 18 years and above in each of rounds three to five of the REACT-2 study, carried out from 15 to 28 September 2020 (round 3), 27 October to 10 November 2020 (round 4) and 25 January to 8 February 2021 (round 5). Registrations closed after ∼190,000 people had signed up at each round.

### Data analysis

We obtained prevalence estimates for reporting of one or more of the 29 symptoms by sex, age and other characteristics, at time of suspected or confirmed COVID-19, and for persistence of symptoms at four and 12 weeks. Our main analyses focused on individual symptoms reported as lasting for 12 weeks (84 days) or more (this excluded a small number of participants with inconsistent or missing data, see Supplementary Figure S1). Prevalence estimates were weighted by sex, age, ethnicity, LTLA population and index of multiple deprivation, to take account of the sampling design that gave approximately equal numbers of participants in each LTLA, and differential response rates, to obtain prevalence estimates that were representative of the population of England as a whole.

We used logistic regression (univariable, and sex, age adjusted) to investigate the associations of demographic and lifestyle factors with persistence of symptoms at 12 weeks or more, and gradient boosted tree models^19^ to investigate predictive ability (area under the curve, AUC) changes from adding variables to the model for persistent symptoms at 12 weeks or more.

To identify a more specific set of persistent symptoms associated with history of COVID-19, in sensitivity analyses, we carried out variable selection in a 30% subset of symptomatic participants: in univariable models, we identified a subset of persistent symptoms (12 or more weeks) that were positively associated with a reported prior positive PCR test, and estimated the population prevalence of persistence of one or more of these symptoms. We also repeated the logistic and gradient boosted tree modeling with this subset of symptoms as outcome variables.

Generalised additive models (GAMs) were constructed with likelihood of symptom persistence at 12 weeks or more modelled as a smoothed function of sex and age. A default thin plate spline was used and the smoothed functions were plotted to visualise the relationship between risk of persistent symptoms and age.

We used free-text analysis to identify single and co-occurring words to indicate other symptoms recorded by participants, and plotted these in a network.

To identify symptom clusters segmenting participants, two binary matrices were constructed for presence or absence (1 or 0) of each of the 29 surveyed symptoms at (i) time of symptom onset and (ii) 12 weeks after, for each participant. Clustering was performed, separately, both row-wise (to identify groups of participants with similar symptoms) and column-wise (to group symptoms based on their co-occurrence) using the CLustering LARge Applications (CLARA) extension of the Partitioning Around Medoids (PAM) algorithm, implemented in the R package *fpc*.^20^ Briefly, PAM searches for the most representative data points to become cluster centroids by minimising the sum of dissimilarities between data points and their assigned centroids. CLARA uses a sampling approach to reduce the computational burden for large data sets. We used Hamming distance as a measure of dissimilarity between participants (row-wise clustering) and symptoms (column-wise clustering). We determined the optimal number of clusters using the average silhouette width. We used two methods to assess cluster stability. First, we bootstrapped and re-clustered 100 times, then quantified the difference between bootstrapped and non-bootstrapped clusters using the Jaccard coefficient, which can range from 0 (no overlap) to 1 (perfect overlap).^21^ Second, we removed each symptom in turn, re-clustered, then calculated the average proportion of non-overlap (APN) between these and whole-dataset clusters as a proxy for the individual variable importance and contribution to the population segmentation.^22^

To further describe patterns of symptom co-occurrence, we took the cross-product of the symptom matrix at symptom onset and at 12 weeks to find pairwise symptom co-occurrence counts, and visualised them as heatmaps.

## Results

### Prevalence of persistent symptoms

A total of 508,707 people took part in REACT-2 rounds three to five and completed surveys. The weighted prevalence of self-reported COVID-19 was 19.2% [19.1,19.3] with 92,116 people reporting one or more of 29 symptoms, of whom 76,155 (82.7%) reported a valid date of symptom onset 12 weeks or more before their survey date (Supplementary Figure S1A). Of those self-reporting COVID-19, 28,713/76,155 (37.7%) experienced at least one symptom for 12 weeks or more and 11,241 (14.8%) experienced at least three symptoms for the same period. This gives a weighted population prevalence of persistent symptoms of 5.75% (5.68, 5.81) for one and 2.22% (2.18, 2.26) for three or more symptoms for England to early February 2021. Almost a third of people with at least one symptom lasting 12 weeks or more (8,771/28,713 [30.5%]) reported having had severe COVID-19 symptoms (“significant effect on my daily life”) at the time of their illness, giving a weighted prevalence overall of people with persistent symptoms at 12 weeks who had reported severe symptoms of 1.72% (1.69,1.76) (Supplementary Table S1A).

Figure 1 shows how the proportion of people with one or multiple symptoms declined over time since infection. There was a rapid drop-off by four weeks, a further, smaller drop by 12 weeks, but then little evidence of further decline over time up to ∼22 weeks for both men and women, with higher prevalence of symptoms at each time point among women.

**Figure 1.**
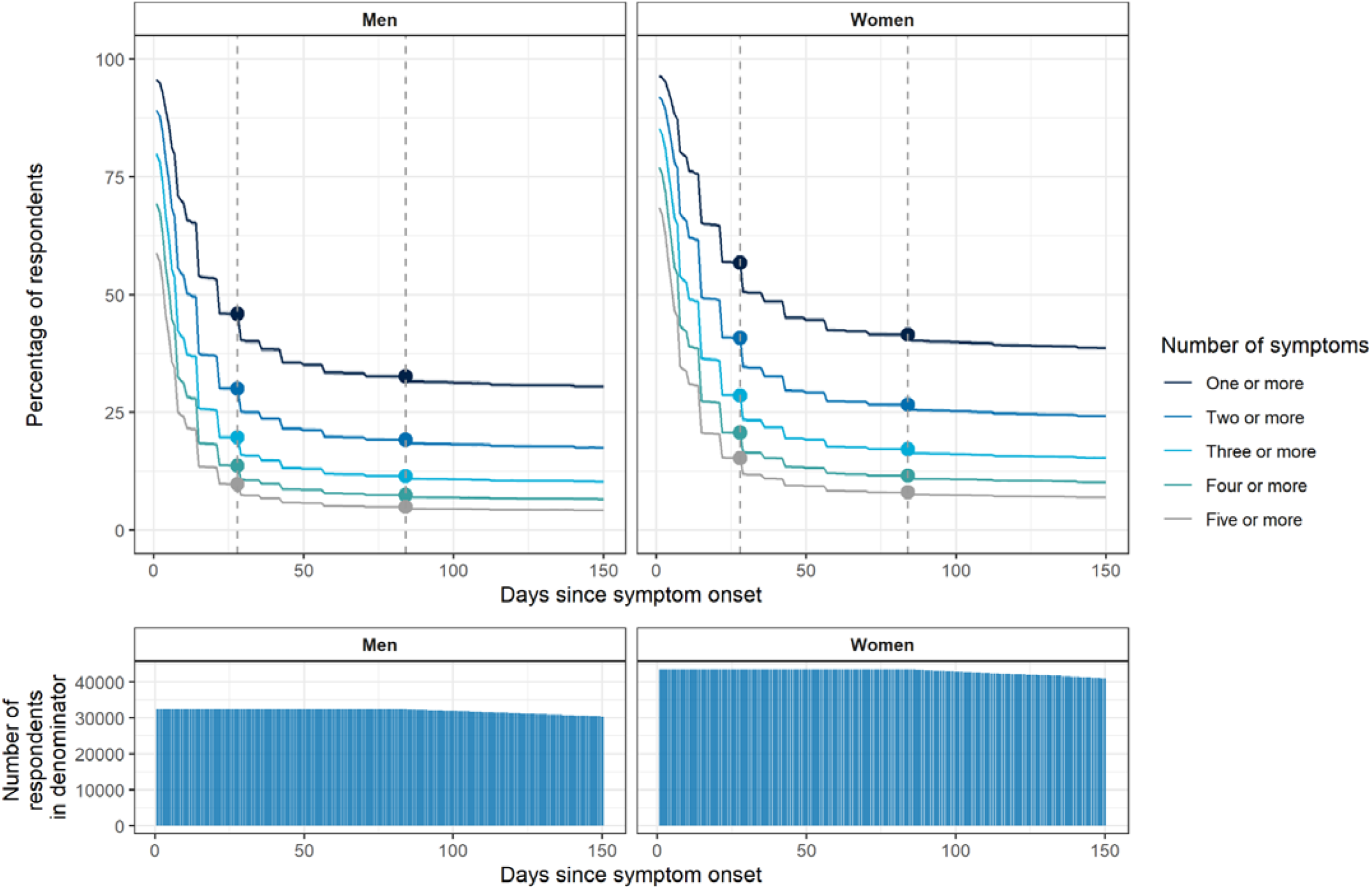
Plots showing persistence of symptoms as a proportion of those who reported symptoms at any time· Plots account for ‘censoring’ – i.e. the denominator in the proportion at day X is the number of respondents who reported a symptomatic infection X or more days prior to responding to the REACT-2 survey (the denominator is shown in the bottom panel plots). Women have higher rates of persistent symptoms; a slower decline in symptom prevalence is observed after 12 weeks in both sexes. The vertical dashed lines show 4 and 12 weeks.

### Factors associated with persistent symptoms

Among symptomatic people, the persistence of one or more symptoms for 12 weeks or more was higher in women than men (age-adjusted OR: 1.51 [1.46,1.55]), and increased with age, with a linear increase of 3.5 percentage points per decade of life (Table 1, Figure 2, Supplementary Figure S2, Supplementary Table S2). With adjustment for sex and age, persistent symptoms were associated with self-reported overweight (OR: 1.16 [1.12, 1.21]) and obesity (OR: 1.53 [1.47,1.59]) compared with normal weight individuals, smoking (OR: 1.35 [1.28,1.41]), vaping (OR: 1.26 [1.18,1.34]), healthcare or care home workers (OR: 1.33 [1.26, 1.41]) and hospitalisation with COVID-19 (OR: 3.46 [2.93,4.09]), while Asian ethnicity (OR: 0.80 [0.74,0.88]) was associated with lower risk of persistent symptoms compared to people of white ethnicity (Table 1, Figure 2, Supplementary Table S2).

**Table 1.** Numbers and proportions of participants who reported one or more symptoms (from a list of 29 surveyed symptoms) of COVID-19 at i) time of symptom onset, ii) 4 weeks post symptom onset, and iii) 12 weeks post symptom onset, among the 76,155 symptomatic participants for whom we have 12 weeks’ follow-up and complete data.

|  |  |  | *4 weeks after initial symptom onset* |  | *12 weeks after initial symptom onset* |  |
| --- | --- | --- | --- | --- | --- | --- |
| Category |  | Symptomatic at infection | Number symptomatic | Percentage symptomatic | Number symptomatic | Percentage symptomatic |
| All participants | - | 76155 | 39737 | 52.2 [51.8-52.5] | 28713 | 37.7 [37.4-38.0] |
| Sex | Women | 43654 | 24812 | 56.8 [56.4-57.3] | 18109 | 41.5 [41.0-41.9] |
|  | Men | 32500 | 14924 | 45.9 [45.4-46.5] | 10604 | 32.6 [32.1-33.1] |
| Age group | 18-24 | 5085 | 2133 | 41.9 [40.6-43.3] | 1534 | 30.2 [28.9-31.4] |
|  | 25-34 | 12245 | 5325 | 43.5 [42.6-44.4] | 3784 | 30.9 [30.1-31.7] |
|  | 35-44 | 15992 | 7646 | 47.8 [47.0-48.6] | 5232 | 32.7 [32.0-33.4] |
|  | 45-54 | 17548 | 9490 | 54.1 [53.3-54.8] | 6858 | 39.1 [38.4-39.8] |
|  | 55-64 | 15148 | 8787 | 58.0 [57.2-58.8] | 6461 | 42.7 [41.9-43.4] |
|  | 65-74 | 7811 | 4823 | 61.7 [60.7-62.8] | 3615 | 46.3 [45.2-47.4] |
|  | 74+ | 2326 | 1533 | 65.9 [64.0-67.8] | 1229 | 52.8 [50.8-54.9] |
| Ethnicity | Asian | 2676 | 1180 | 44.1 [42.2-46.0] | 809 | 30.2 [28.5-32.0] |
|  | Black | 739 | 361 | 48.8 [45.3-52.5] | 278 | 37.6 [34.2-41.2] |
|  | Mixed | 1244 | 631 | 50.7 [47.9-53.5] | 487 | 39.1 [36.5-41.9] |
|  | Other | 793 | 413 | 52.1 [48.6-55.5] | 299 | 37.7 [34.4-41.1] |
|  | White | 70103 | 36838 | 52.5 [52.2-52.9] | 26603 | 37.9 [37.6-38.3] |
| BMI | Normal weight | 26128 | 12803 | 49.0 [48.4-49.6] | 8954 | 34.3 [33.7-34.8] |
|  | Obese | 16076 | 9416 | 58.6 [57.8-59.3] | 7107 | 44.2 [43.4-45.0] |
|  | Overweight | 23958 | 12429 | 51.9 [51.2-52.5] | 8916 | 37.2 [36.6-37.8] |
|  | Underweight | 886 | 450 | 50.8 [47.5-54.1] | 333 | 37.6 [34.5-40.8] |
| Healthcare or care home worker | Yes | 5243 | 3041 | 58.0 [56.7-59.3] | 2283 | 43.5 [42.2-44.9] |
| Index of multiple deprivation (IMD) quintile | 1 - most deprived | 8096 | 4438 | 54.8 [53.7-55.9] | 3445 | 42.6 [41.5-43.6] |
|  | 2 | 12789 | 6836 | 53.5 [52.6-54.3] | 5066 | 39.6 [38.8-40.5] |
|  | 3 | 16491 | 8722 | 52.9 [52.1-53.7] | 6336 | 38.4 [37.7-39.2] |
|  | 4 | 18202 | 9529 | 52.4 [51.6-53.1] | 6728 | 37.0 [36.3-37.7] |
|  | 5 - least deprived | 20577 | 10212 | 49.6 [48.9-50.3] | 7138 | 34.7 [34.0-35.3] |
| Current smoker | Current cigarette smoker | 8441 | 4536 | 53.7 [52.7-54.8] | 3552 | 42.1 [41.0-43.1] |
|  | Not current cigarette smoker | 67021 | 34880 | 52.0 [51.7-52.4] | 24916 | 37.2 [36.8-37.5] |
|  | Prefer not to say | 693 | 321 | 46.3 [42.6-50.0] | 245 | 35.4 [31.9-39.0] |
| Current vaper | Current vaper | 4933 | 2534 | 51.4 [50.0-52.8] | 1986 | 40.3 [38.9-41.6] |
|  | Not current vaper | 70802 | 37015 | 52.3 [51.9-52.6] | 26586 | 37.5 [37.2-37.9] |
|  | Prefer not to say | 420 | 188 | 44.8 [40.1-49.5] | 141 | 33.6 [29.2-38.2] |
| Severity of COVID symptoms | Mild symptoms | 21454 | 7519 | 35.0 [34.4-35.7] | 5441 | 25.4 [24.8-25.9] |
|  | Moderate symptoms | 37976 | 20413 | 53.8 [53.3-54.3] | 14501 | 38.2 [37.7-38.7] |
|  | Severe symptoms | 16725 | 11805 | 70.6 [69.9-71.3] | 8771 | 52.4 [51.7-53.2] |
| Treatment sought/ received for COVID | No medical attention sought | 47642 | 21009 | 44.1 [43.7-44.5] | 15098 | 31.7 [31.3-32.1] |
|  | Sought medical attention from pharmacist / by phone (NHS 111/GP) | 17794 | 11713 | 65.8 [65.1-66.5] | 8717 | 49.0 [48.3-49.7] |
|  | Visited GP/walk-in centre/A&E | 6154 | 4269 | 69.4 [68.2-70.5] | 2811 | 45.7 [44.4-46.9] |
|  | Hospital admission | 629 | 494 | 78.5 [75.2-81.6] | 408 | 64.9 [61.1-68.5] |
| Gross household income | 0-14,999 | 4543 | 2862 | 63.0 [61.6-64.4] | 2316 | 51.0 [49.5-52.4] |
|  | 15,000-49,999 | 21468 | 11847 | 55.2 [54.5-55.8] | 8746 | 40.7 [40.1-41.4] |
|  | 50,000-149,999 | 21923 | 10382 | 47.4 [46.7-48.0] | 7100 | 32.4 [31.8-33.0] |
|  | >150,000 | 3082 | 1330 | 43.2 [41.4-44.9] | 886 | 28.7 [27.2-30.4] |
| Comorbidities | Organ transplant recipient | 70 | 43 | 61.4 [49.7-72.0] | 35 | 50.0 [38.6-61.4] |
|  | Diabetes (type I or II) | 3282 | 2045 | 62.3 [60.6-64.0] | 1596 | 48.6 [46.9-50.3] |
|  | Heart disease or heart problems | 2822 | 1894 | 67.1 [65.4-68.8] | 1521 | 53.9 [52.1-55.7] |
|  | Hypertension (high blood pressure) | 8269 | 5203 | 62.9 [61.9-64.0] | 4009 | 48.5 [47.4-49.6] |
|  | Stroke | 415 | 261 | 62.9 [58.1-67.4] | 211 | 50.8 [46.0-55.6] |
|  | Kidney disease | 547 | 405 | 74.0 [70.2-77.5] | 325 | 59.4 [55.2-63.5] |
|  | Liver disease | 309 | 209 | 67.6 [62.2-72.6] | 171 | 55.3 [49.8-60.8] |
|  | Anaemia | 1371 | 952 | 69.4 [66.9-71.8] | 801 | 58.4 [55.8-61.0] |
|  | Asthma | 9119 | 5616 | 61.6 [60.6-62.6] | 4419 | 48.5 [47.4-49.5] |
|  | Other lung condition | 1639 | 1252 | 76.4 [74.3-78.4] | 1113 | 67.9 [65.6-70.1] |
|  | Cancer | 820 | 533 | 65.0 [61.7-68.2] | 411 | 50.1 [46.7-53.5] |
|  | Condition affecting the brain and nerves | 686 | 457 | 66.6 [63.0-70.0] | 378 | 55.1 [51.4-58.8] |
|  | A weakened immune system/reduced ability to deal with infections | 3080 | 2165 | 70.3 [68.7-71.9] | 1815 | 58.9 [57.2-60.7] |
|  | Depression | 6911 | 4680 | 67.7 [66.6-68.8] | 3931 | 56.9 [55.7-58.0] |
|  | Anxiety | 10516 | 6768 | 64.4 [63.4-65.3] | 5553 | 52.8 [51.9-53.8] |
|  | Psychiatric disorder | 641 | 417 | 65.1 [61.3-68.6] | 372 | 58.0 [54.2-61.8] |

**Figure 2.**
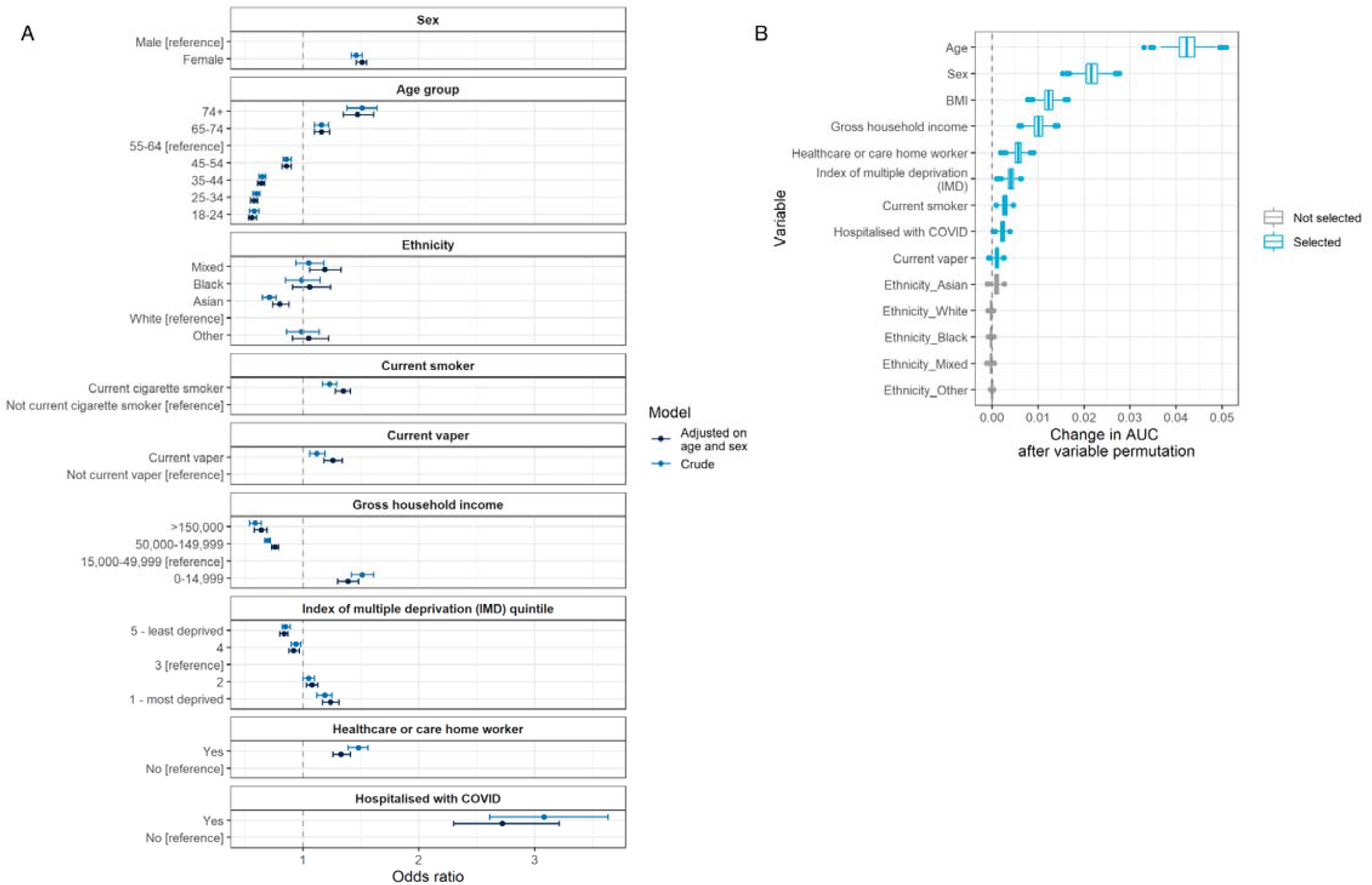
(A) Logistic regression models with one or more symptoms at 12 weeks (y/n) as the binary outcome variable, both unadjsuted and adjusted for sex and age; (B) Mean contribution to area under the curve (AUC) that each variable makes to a multivariable boosted tree model, derived by permuting each variable in turn (1000x to obtain a distribution) and quantifying the change in model performance. Age, sex, body mass index (BMI), household income, healthcare/care home worker, deprivation, smoking status and prior hospitalisation with COVID-19 are the strongest predictors of persistent symptoms in multivariable modelling, while Asian ethnicity is associated with a lower risk of persistent symptoms at 12 weeks.

There was a higher proportion with persistent symptoms among those with low incomes at 51.0% (49.5, 52.4) compared with high incomes at 28.7% (27.2, 30.4) and among people living in the most deprived areas at 42.6% (41.5, 43.6) compared with the most affluent areas at 34.7% (34.0, 35.3) (Table 1).

Prevalence of persistent symptoms at 12 or more weeks was around 50% or more among people reporting co-morbidities, ranging up to 67.9% (65.6,70.1) for “other lung condition” (Table 1).

In addition to the 29 symptoms enquired about on the questionnaire, 8,370 respondents gave free-text descriptions of other symptoms, of whom 1,860 reported symptoms that persisted for 12 weeks or more. Free-text analysis of co-occurring words indicated common additional symptoms which were not in our survey, including brain-fog, hair-loss, blood-pressure, heart-palpitations, severe-joint-pain (Supplementary Figure S3).

### Sensitivity analysis

In sensitivity analyses, we assessed the extent by which the observed prevalence of, and factors associated with, persistent symptoms were altered by using a smaller set of symptoms. To improve specificity of the symptoms for COVID-19, we selected symptoms that were positively predictive of self-reported PCR positivity (among a 30% holdout set of people, Supplementary Figure S1), thus identifying a subset of 15 of the surveyed 29 symptoms (Supplementary Table S1B, Supplementary Figure S4). Based on this more limited set, the number of people with symptoms reporting one or more persistent symptoms at 12 weeks was 24,867/76,155 (32.7%), giving a weighted population prevalence of 4.95% (4.89, 5.01) (Supplementary Table S1B).

Logistic regression modelling with persistence of any one of these 15 symptoms at 12 weeks as a binary outcome showed similar results to the full 29-symptom analysis (Figure 2, Supplementary Figure S5).

### Clustering analysis

In clustering analysis, two stable clusters of participants were identified based on symptom profiles at 12 weeks. Participants in Cluster L1 (“tiredness cluster”) experienced high prevalence of tiredness, which co-occurred with muscle aches, difficulty sleeping and shortness of breath (Supplementary Figure S6). Participants in Cluster L2 (“respiratory cluster”) experienced high prevalence of respiratory symptoms including shortness of breath and tight chest, as well as chest pain (Figure 3, Figure 4). A higher proportion of people in the respiratory cluster reported severe symptoms at the time of their COVID-19 illness (43.5%, [42.0,44.9]) than in the tiredness cluster (27.4%, [26.7,28.1]) (Supplementary Table S3).

**Figure 3.**
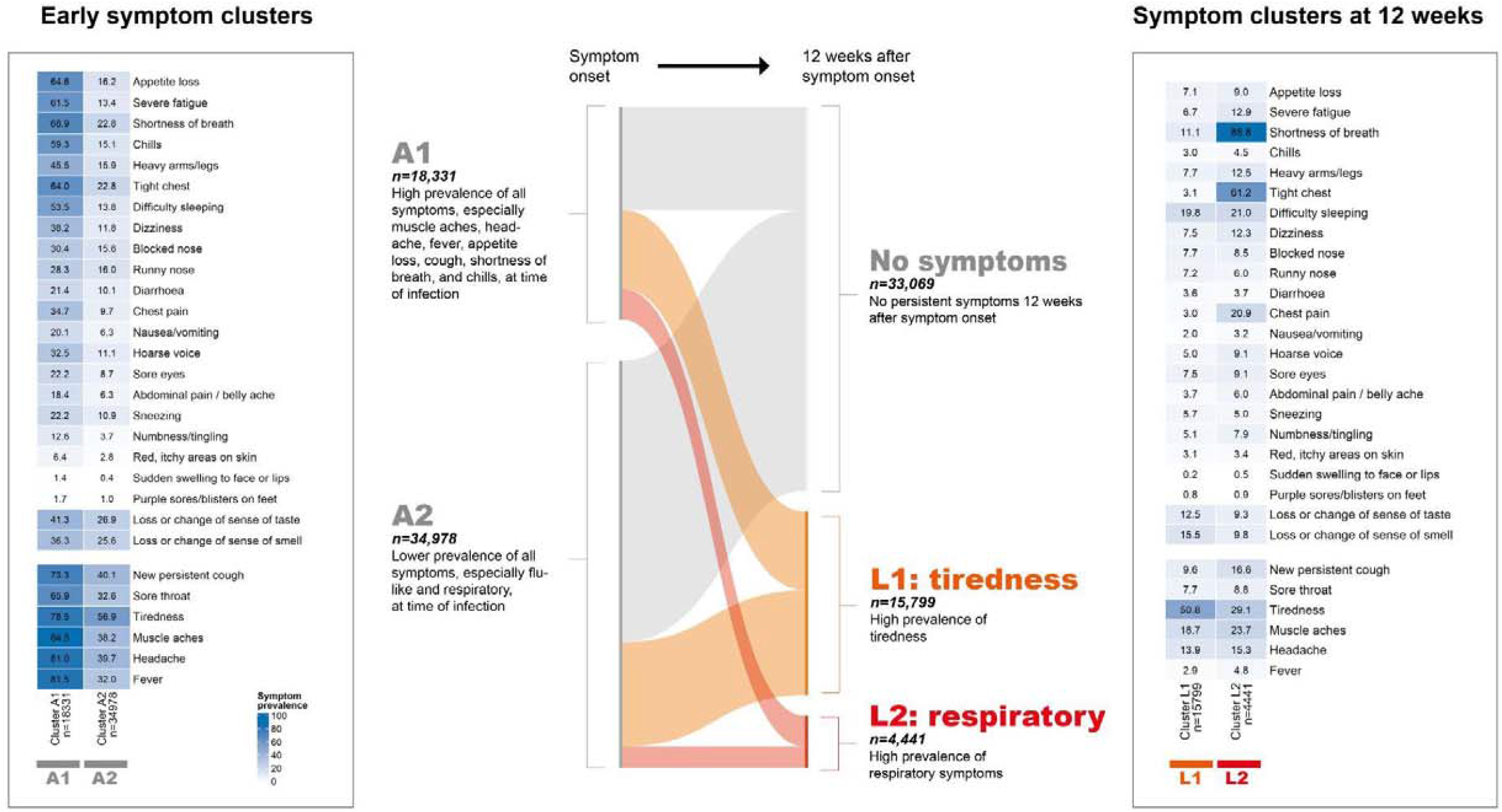
Results of clustering on symptom profile at time of symptom onset and then reclustering at 12 weeks, using CLARA (partitioning around medoids) algorithm. Central Sankey plot shows transitions between early and 12-week clusters. Two stable clusters of symptomatic infections were identified at t0: cluster A1 was characterised primarily by higher prevalence of flu-like symptoms (muscle aches, headache, fever, appetite loss, chills) and respiratory symptoms (shortness of breath, tight chest, new persistent cough). Two stable clusters were identified at 12 weeks. Cluster L1 (“tiredness cluster”) had high prevalence of tiredness. Cluster L2 (“respiratory cluster”) was a smaller subset of 4,441 participants who had high prevalence of shortness of breath and tight chest as well as chest pain.

**Figure 4.**
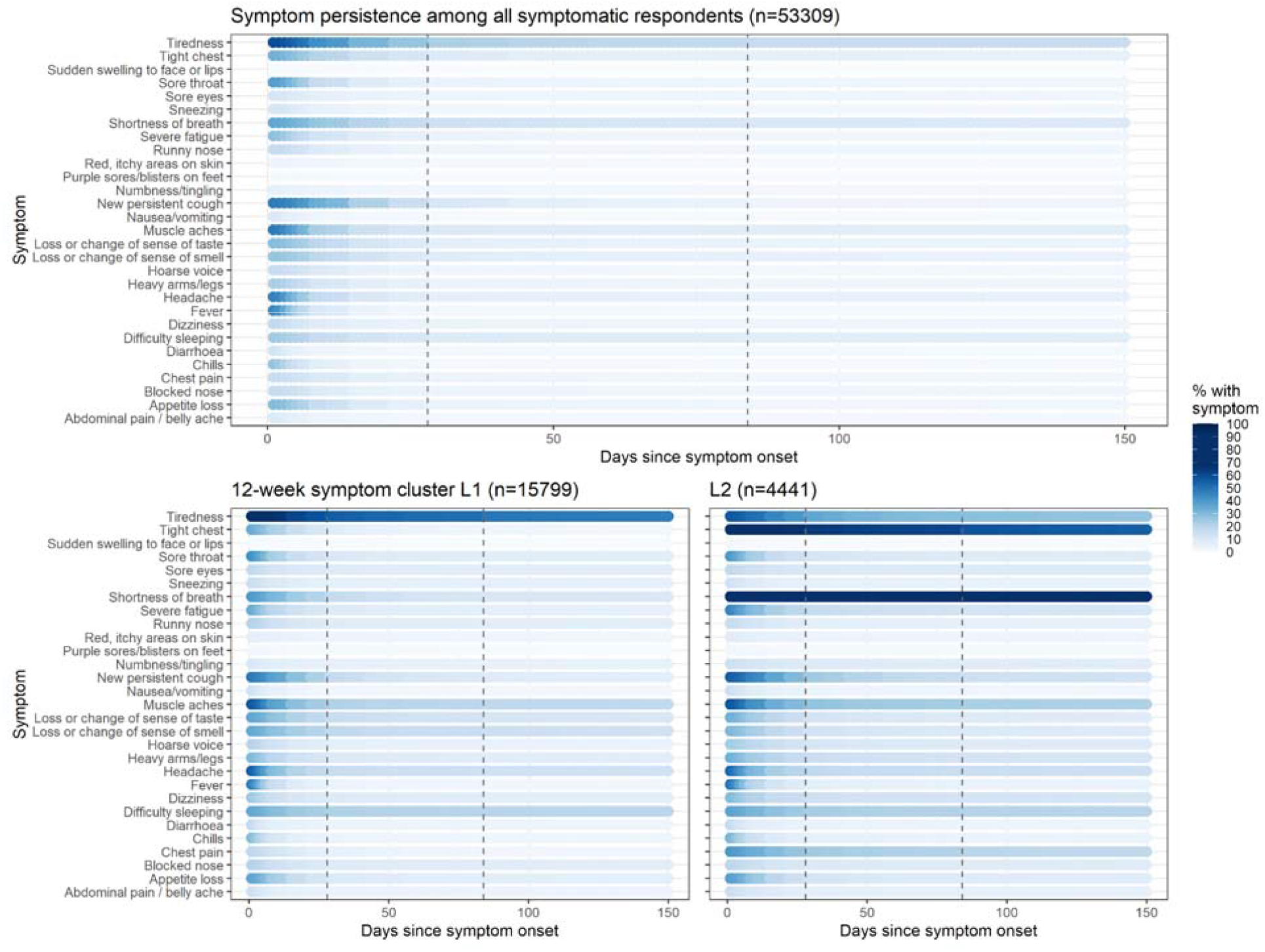
Persistence of symptoms for all symptomatic participants (top panel) and in 12-week symptoms clusters L1 and L2 (bottom panels). Dashed lines show 4 and 12 weeks post symptom onset.

We obtained research ethics approval from the South Central-Berkshire B Research Ethics Committee (IRAS ID: 283787). The REACT Public Advisory Panel provides regular review of the study processes and results.

## Discussion

In this large community-based study of symptoms following COVID-19 among adults aged 18 years and above in England, participants reported high prevalence of persistent symptoms lasting 12 weeks or more. Estimates ranged from 5.8% of the population experiencing one or more persistent symptoms post-COVID-19 (corresponding to over 2 million adults in England), to 2.2% for three or more persistent symptoms (just under a million adults in England), and 1.7% with one or more symptoms lasting at least 12 weeks in people who reported severe COVID-19 symptoms affecting their daily life at the time of their illness.

Our estimates of the proportion of people with persistent COVID-19 symptoms are higher than in many other studies, although previous estimates have varied widely. At the low end, one study found that 2.3% of people with COVID-19 still reported symptoms at 12 weeks;^6^ other studies have reported 13.7% of people were symptomatic at 84 days,^7^ 14.8% symptomatic at 90 days,^8^ 27% at 60 days,^23^ 34.7% at 7 months,^24^ and as high as 46% at six months.^9^ Our comparatively high estimate, at 37.7% of people with COVID-19 experiencing one or more symptoms at 12 weeks, may partly reflect the large list of symptoms we surveyed, many of which are common and not specific to COVID-19. However, we asked participants only about symptoms that they related to a confirmed or suspected episode of COVID-19, and not to symptoms more generally. In addition, to improve specificity, in a sensitivity analysis we restricted COVID-19 symptoms to a subset of 15 that were predictive of reported PCR positivity and which included among others, loss or change of sense of smell or taste, tiredness, shortness of breath, muscle aches, heavy arms/legs, severe fatigue, tight chest and chest pain. This gave a similar proportion of people with persistent symptoms at 12 weeks (32.7%), and a population prevalence (5%) similar to that obtained when all 29 surveyed symptoms were considered.

Increasing age, female sex, BMI, hospitalisation and co-morbidites have previously been identified as risk factors for Long COVID^6,25^. Our finding of a linear association between age and persistent symptoms in people with symptomatic COVID-19 contrasts with some other studies that suggest the highest prevalence is found in middle-aged groups.^7^ Our finding is conditional on symptomatic COVID-19, reflecting the fact that older age groups in the community have lower infection rates than younger people^26^ and are more likely to be asymptomatic.^27^ Our findings of an association of persistent symptoms with deprivation, and with smoking or vaping, are not well described in the literature, although smokers have been found to have a higher risk of severe COVID-19.^28^

Our identification of two stable and well-differentiated symptom clusters at 12 weeks supports the characterisation of Long COVID as a diverse set of overlapping conditions. Previous studies have taken a similar unsupervised approach to characterising subtypes of Long COVID, albeit at earlier time points: Sudre et al.^6^ identified two symptom clusters at 28 days post-symptom-onset, which resembled the early clusters identified in our study but not the 12-week clusters. Huang et al.^23^ identified five clusters at 61 days, two of which reported high prevalences of respiratory symptoms similar to cluster L2 in our study.

### Strengths and Limitations

This study uses a large random community sample with a high response rate (26–29% across the three rounds) to describe the persistence of COVID-19 symptoms. It is therefore more likely to be representative of the range of disease severity in the population compared to some others, especially those based on hospitalised cases alone^5^. The focus in our questionnaire on persistence of self-reported COVID-19 symptoms, without specific reference to Long COVID (in contrast to the questions in some other studies^7^) has allowed us to investigate the persistence of a wide range of specific symptoms that have been suggested as relevant to Long COVID.^3,5^ However, it is clear that a wide spectrum of symptoms and clinical presentations post-COVID-19 may be involved; our open free-text question identified a number of symptoms not included in our questionnaire including “brain fog”, “palpitations” and “hair loss”.^29^ However, as the study was based on self-reported data and because many of the symptoms are common and not specific to COVID-19, we may, as noted, have overestimated the prevalence of persistent symptoms.

A further limitation is the retrospective study design, which introduces the possibility of recall bias. Nonetheless, in earlier analyses we have shown that participant reports of date of onset of their symptoms produce an epidemic curve that very closely tracks the epidemic.^27,30,31^ Respondents were restricted to reporting a single date of (initial) symptom onset which does not allow for delayed onset of some symptoms, nor does it allow for the reporting of relapsing symptoms which appear to be a feature of Long COVID.^6^ A further limitation, despite the high response rate for a community surveillance study, is the possibility of participation bias as the REACT-2 study included a home antibody self-test;^27^ it is plausible that people with persistent symptoms may have been more likely to participate in order to ascertain their antibody status.

### Implications

The scale of morbidity identified in this study presents significant challenges for the affected individuals and their families, and for health services and society more broadly. After the initial decline in symptom prevalence between 4 and 12 weeks the prevalence of persistent symptoms plateaued indicating that large numbers of people may have chronic symptoms requiring investigation and intervention including rehabilitation. We show here that economically disadvantaged people and those in deprived areas appear to have a higher burden of persistent symptoms post COVID-19, compounding the excess burden of severe illness and mortality from COVID-19 experienced by these groups^32^. Inability to work due to disability from Long COVID could have a high impact on these populations. Given the high potential population health burden, investment is urgently needed to expand the network of services investigating and managing people with Long COVID, to link these services to national research studies and consistent data collection to improve our understanding, and to create the infrastructure for trials in the same way as has been achieved for acute COVID-19.

We identified two clusters of participants based on their symptoms. Individuals with predominantly respiratory symptoms comprised around one quarter of those with persistent symptoms --the larger proportion comes from a cluster of less organ-specific symptoms, particularly fatigue. Clinicians will need to be aware of the range of presenting symptoms to best support their patients towards recovery and there will need to be education of healthcare professionals to recognise and respect the experiences of people reporting these symptoms. Societal recognition of Long COVID as an important outcome of the pandemic (including as an occupational illness for those acquiring it through work) is needed, including for those who may require benefits as well as rehabilitation. The Long COVID patient groups provide much needed support to those involved, but will require resources if they are to be able to work effectively with health services, especially to reach people in more deprived and marginalised groups.

In summary we have identified significant ongoing morbidity among people post COVID-19, with a substantial proportion experiencing persistent symptoms lasting 12 weeks or more. Managing the long term sequelae of SARS-CoV-2 infection in the population will remain a major challenge for health services in the next stage of the pandemic.

## Data Availability

Tables with summary data used in this report are available here: https://github.com/mrc-ide/reactidd/tree/master/inst/extdata/react2_long_COVID_paper

## Data availability

Summary tabular data are provided with this paper.

## Declaration of interests

We declare no competing interests.

## Author contributions

MW -conceptualisation, formal analysis, visualisation, methodology, writing – original draft, writing– review & editing; JE -conceptualisation, formal analysis, methodology, writing – original draft, writing– review & editing; MCH -supervision, methodology, writing– review & editing; SR: supervision, methodology, writing– review & editing; AD -funding acquisition, supervision, writing– review & editing; GC -conceptualisation, supervision, methodology, writing – original draft, writing– review & editing; HW -conceptualisation, supervision, methodology, writing – original draft, writing– review & editing; PE -funding acquisition, conceptualisation, supervision, methodology, writing – original draft, writing– review & editing.

## Acknowledgements

GC is supported by an NIHR Professorship. WSB is the Action Medical Research Professor, AD is an NIHR senior investigator and DA and PE are Emeritus NIHR Senior Investigators. SR acknowledges support from MRC Centre for Global Infectious Disease Analysis, NIHR Health Protection Research Unit, Wellcome Trust (200861/Z/16/Z, 200187/Z/15/Z), and Centres for Disease Control and Prevention (US, U01CK0005-01-02). HW is a National Institute for Health Research (NIHR) Senior Investigator and acknowledges support from NIHR Biomedical Research Centre of Imperial College NHS Trust, NIHR School of Public Health Research, NIHR Applied Research Collaborative North West London, and Wellcome Trust (UNS32973). PE is Director of the MRC Centre for Environment and Health (MR/L01341X/1, MR/S019669/1). PE acknowledges support from the NIHR Imperial Biomedical Research Centre and the NIHR HPRUs in Chemical and Radiation Threats and Hazards and in Environmental Exposures and Health, the British Heart Foundation Centre for Research Excellence at Imperial College London (RE/18/4/34215), Health Data Research UK (HDR UK) and the UK Dementia Research Institute at Imperial (MC_PC_17114). We thank The Huo Family Foundation for their support of our work on COVID-19. Our work on Long COVID is also being supported by grants from NIHR and UK Research and Innovation (UKRI): REACT GE (MR/V030841/1) and COV-LT-0040 – REACT Long COVID (REACT-LC).

We thank key collaborators on this work --Ipsos MORI: Stephen Finlay, John Kennedy, Kevin Pickering, Duncan Peskett, Sam Clemens and Kelly Beaver; Institute of Global Health Innovation at Imperial College London: Gianluca Fontana, Sutha Satkunarajah, Didi Thompson; the Imperial Patient Experience Research Centre and the REACT Public Advisory Panel; NHS Digital for access to the NHS Register; Dr Nisreen Alwan.

## Supplementary material

**Table S1.**
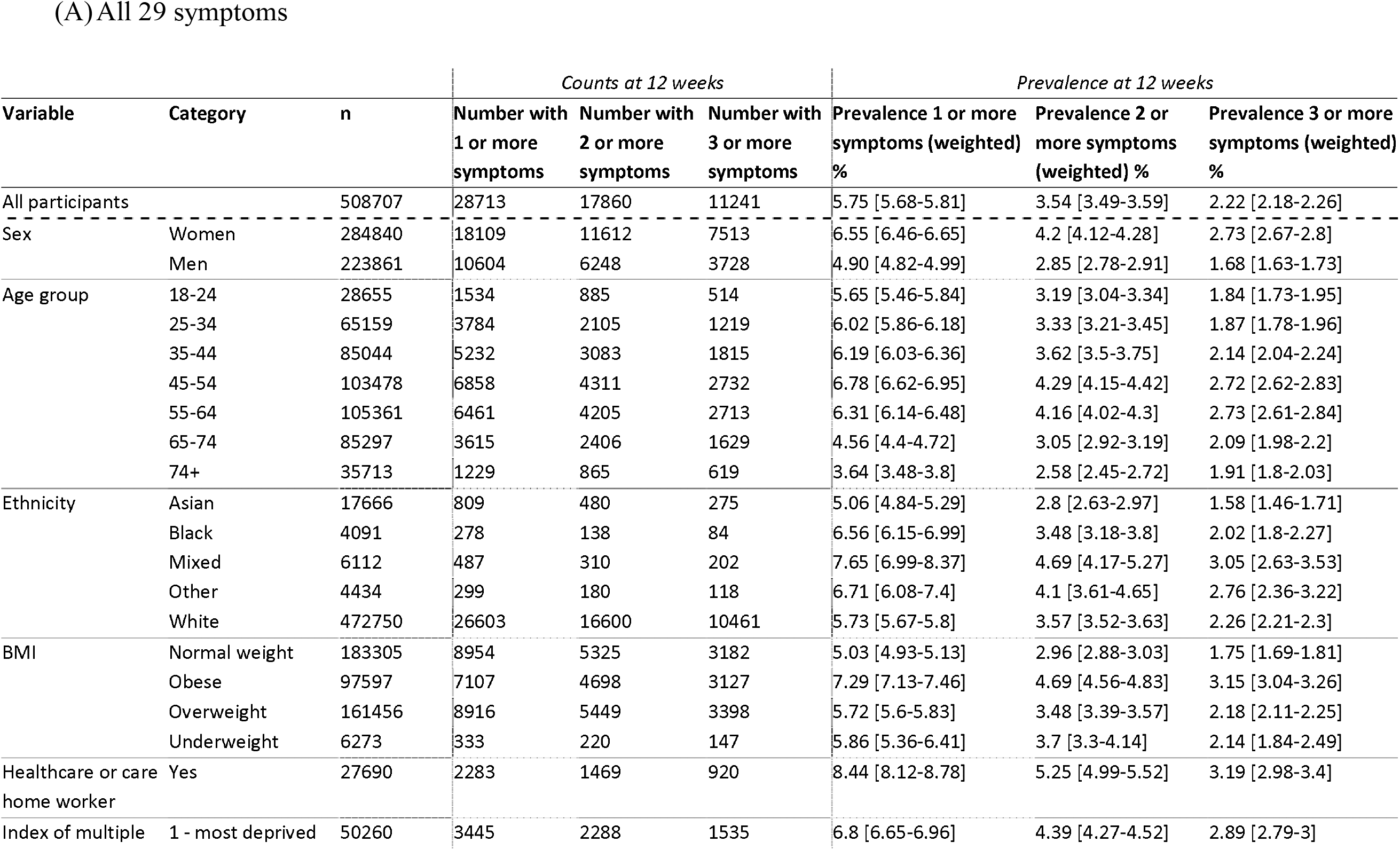
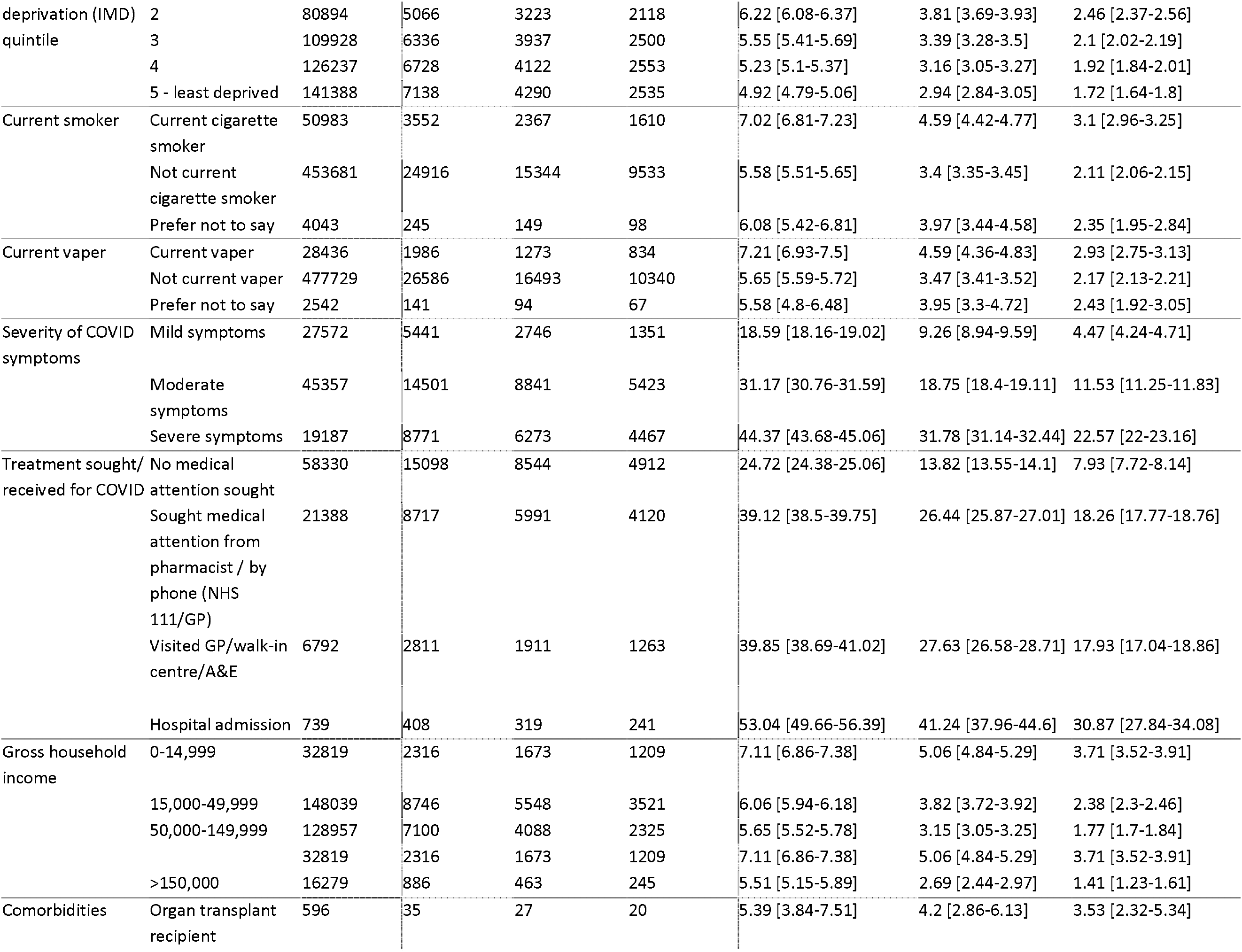
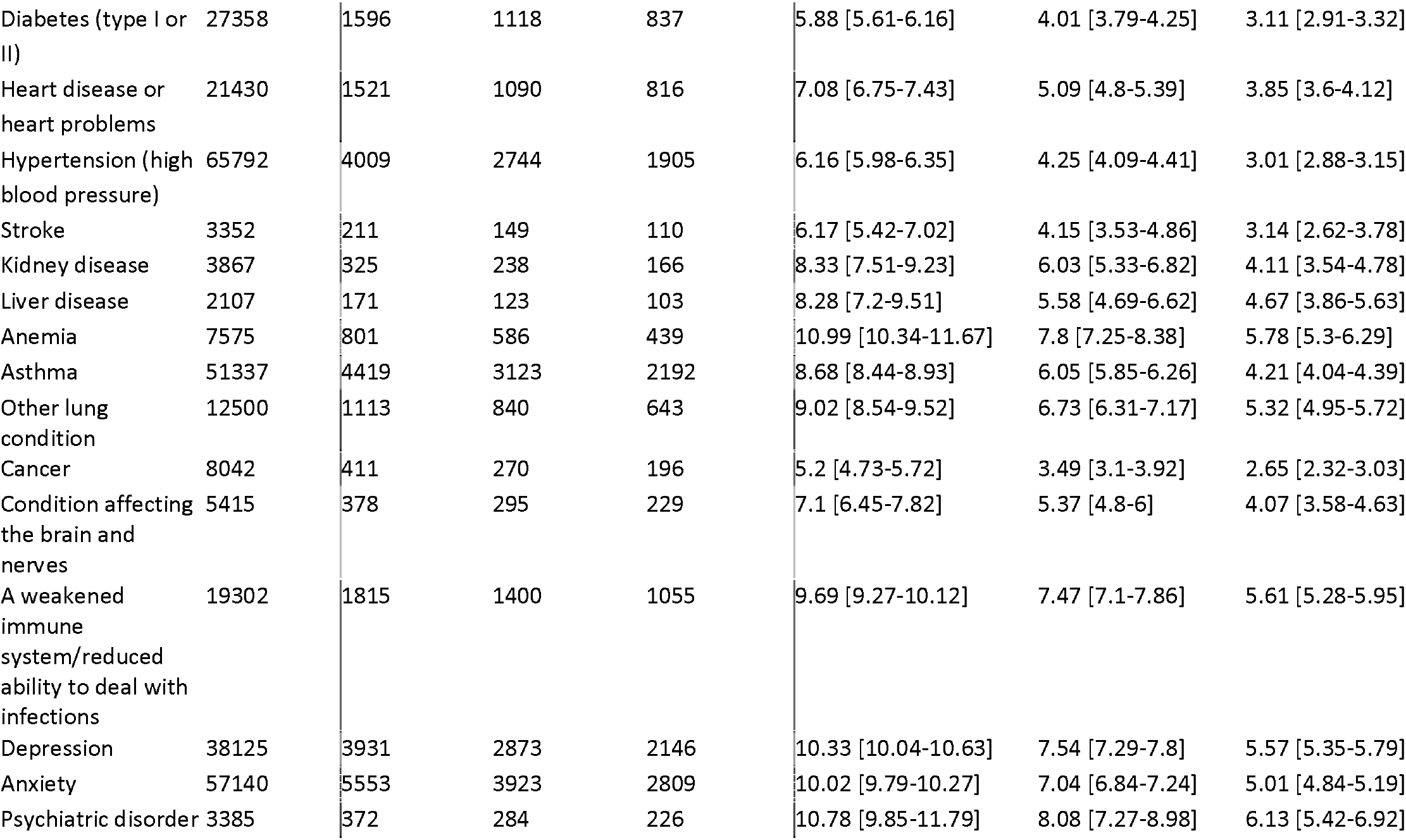
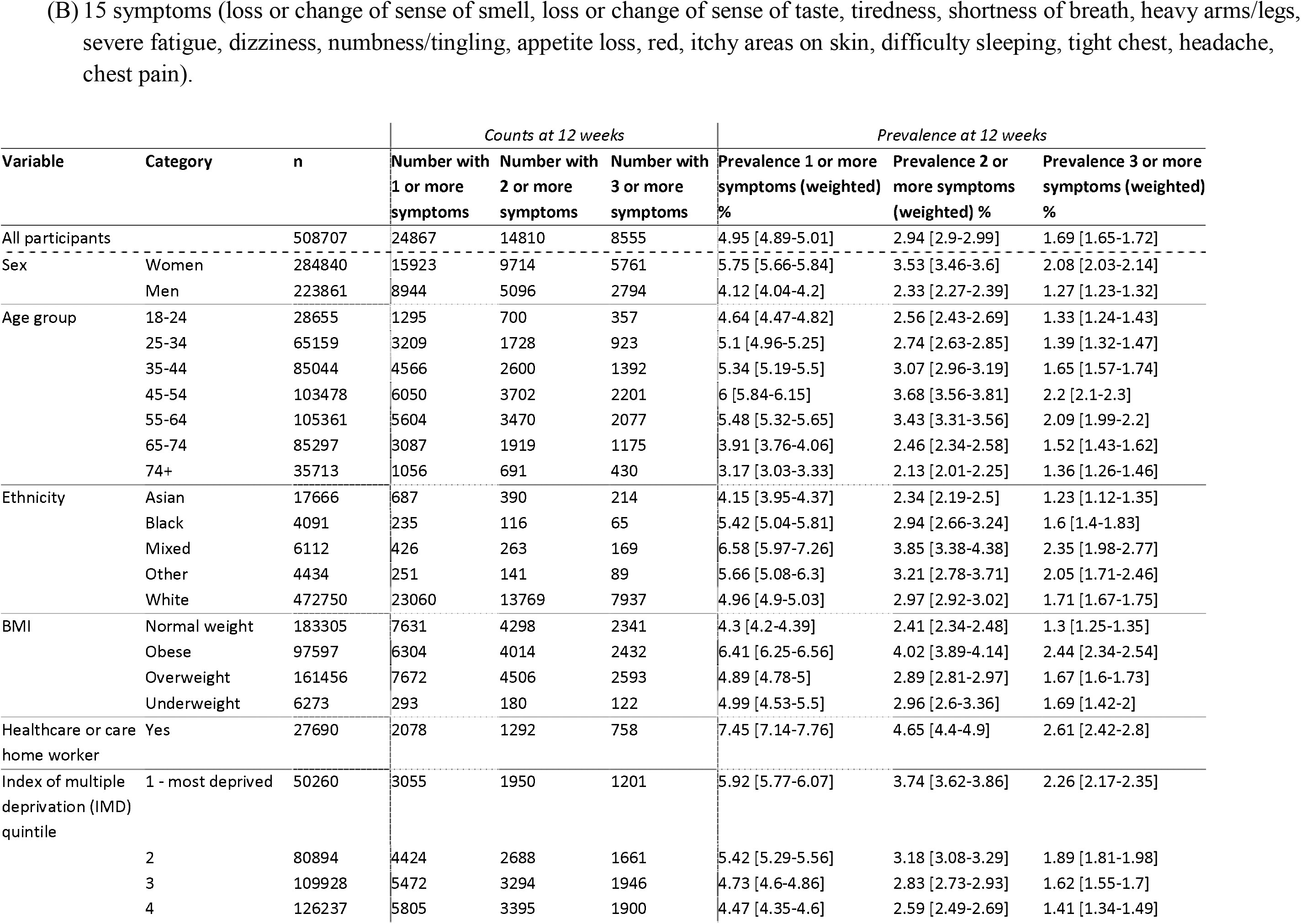
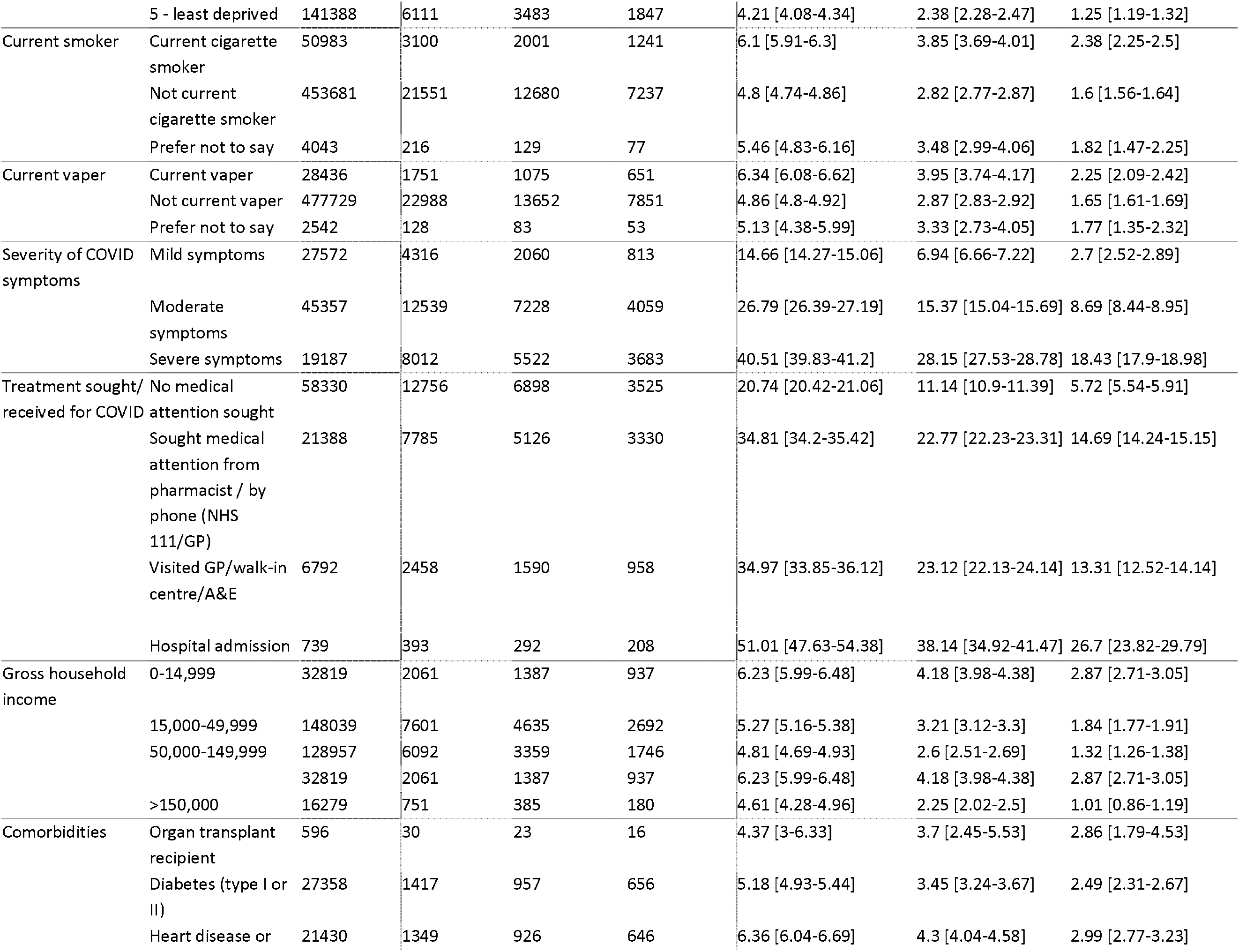
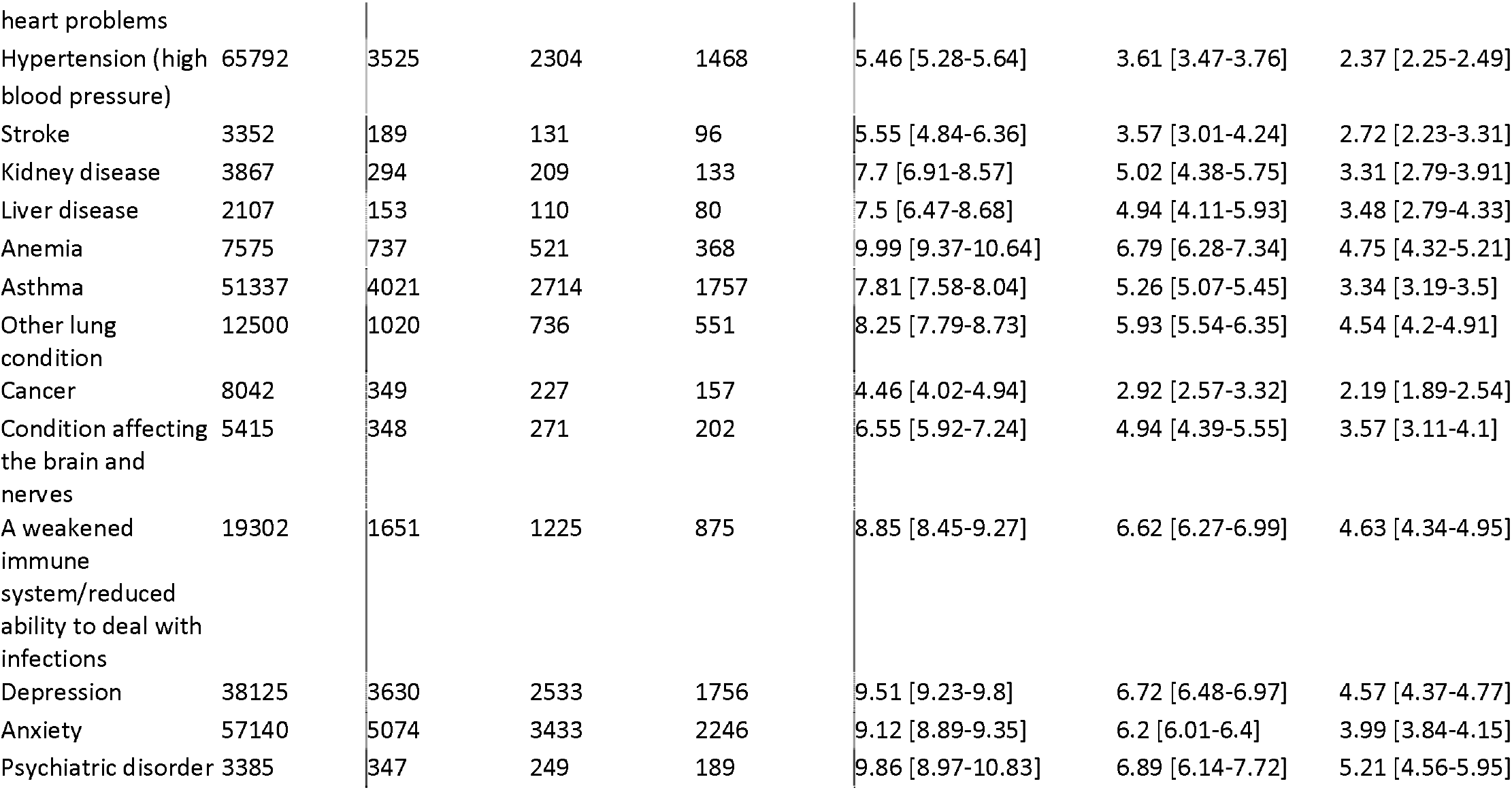
Numbers and weighted prevalence of currently or previously reported persistent symptoms in the full REACT-2 study population (rounds 3–5). Table 1A shows prevalences of any of the 29 surveyed symptoms; Table 1B shows prevalences of any of a subset of 15 symptoms identified as predictive of PCR positivity. The final three columns show prevalences weighted to be representative of the adult population of England, by age, sex, ethnicity, lower-tier local authority population, and index of multiple deprivation (IMD).

**Table S2.** Odds ratios for persistent symptoms at 12 weeks among symptomatic respondents, derived from logistic regression models (forest plot shown in Figure 2)

| Predictor | Level | Crude (univariate) OR | Adjusted on age and sex |
| --- | --- | --- | --- |
| Sex | Male [reference] | - | - |
|  | Female | 1.46 [1.42-1.51] | 1.51 [1.46-1.55] |
| Age group | 18-24 | 0.58 [0.54-0.62] | 0.56 [0.53-0.6] |
|  | 25-34 | 0.6 [0.57-0.63] | 0.58 [0.55-0.61] |
|  | 35-44 | 0.65 [0.62-0.68] | 0.64 [0.61-0.67] |
|  | 45-54 | 0.86 [0.83-0.9] | 0.86 [0.82-0.9] |
|  | 55-64 [reference] | - | - |
|  | 65-74 | 1.16 [1.1-1.22] | 1.16 [1.1-1.23] |
|  | 74+ | 1.51 [1.38-1.64] | 1.47 [1.35-1.61] |
| Ethnicity | White [reference] | - | - |
|  | Asian | 0.71 [0.65-0.77] | 0.8 [0.74-0.88] |
|  | Black | 0.99 [0.85-1.15] | 1.06 [0.91-1.24] |
|  | Mixed | 1.05 [0.94-1.18] | 1.19 [1.06-1.33] |
|  | Other | 0.99 [0.86-1.14] | 1.05 [0.91-1.22] |
| BMI | Obese | 1.52 [1.46-1.58] | 1.53 [1.47-1.59] |
|  | Overweight | 1.14 [1.1-1.18] | 1.16 [1.12-1.21] |
|  | Normal weight [reference] | - | - |
|  | Underweight | 1.15 [1.01-1.33] | 1.17 [1.02-1.35] |
| Healthcare or care home worker | No [reference] | - | - |
|  | Yes | 1.48 [1.39-1.56] | 1.33 [1.26-1.41] |
| Index of multiple deprivation (IMD) quintile | 1 - most deprived | 1.19 [1.12-1.25] | 1.24 [1.17-1.31] |
|  | 2 | 1.05 [1-1.1] | 1.08 [1.03-1.13] |
|  | 3 [reference] | - | - |
|  | 4 | 0.94 [0.9-0.98] | 0.92 [0.88-0.97] |
|  | 5 - least deprived | 0.85 [0.82-0.89] | 0.84 [0.8-0.87] |
| Current smoker | Not current cigarette smoker [reference] | - | - |
|  | Current cigarette smoker | 1.23 [1.17-1.29] | 1.35 [1.28-1.41] |
|  | Prefer not to say | 0.92 [0.79-1.08] | 1.03 [0.88-1.21] |
| Current vaper | Not current vaper [reference] | - | - |
|  | Current vaper | 1.12 [1.06-1.19] | 1.26 [1.18-1.34] |
|  | Prefer not to say | 0.84 [0.69-1.03] | 0.94 [0.77-1.16] |
| Treatment sought/ received for COVID | No medical attention sought [reference] | - | - |
|  | Hospital admission | 3.98 [3.37-4.69] | 3.46 [2.93-4.09] |
|  | Sought medical attention from pharmacist / by phone (NHS 111/GP) | 2.07 [2-2.14] | 2.05 [1.98-2.12] |
|  | Visited GP/walk-in centre/A&E | 1.81 [1.72-1.91] | 1.67 [1.58-1.76] |
| Gross household income | 0-14,999 | 1.51 [1.42-1.61] | 1.39 [1.3-1.48] |
|  | 15,000-49,999 [reference] | - | - |
|  | 50,000-149,999 | 0.7 [0.67-0.72] | 0.76 [0.73-0.79] |
|  | >150,000 | 0.59 [0.54-0.64] | 0.64 [0.58-0.69] |

**Table S3.**
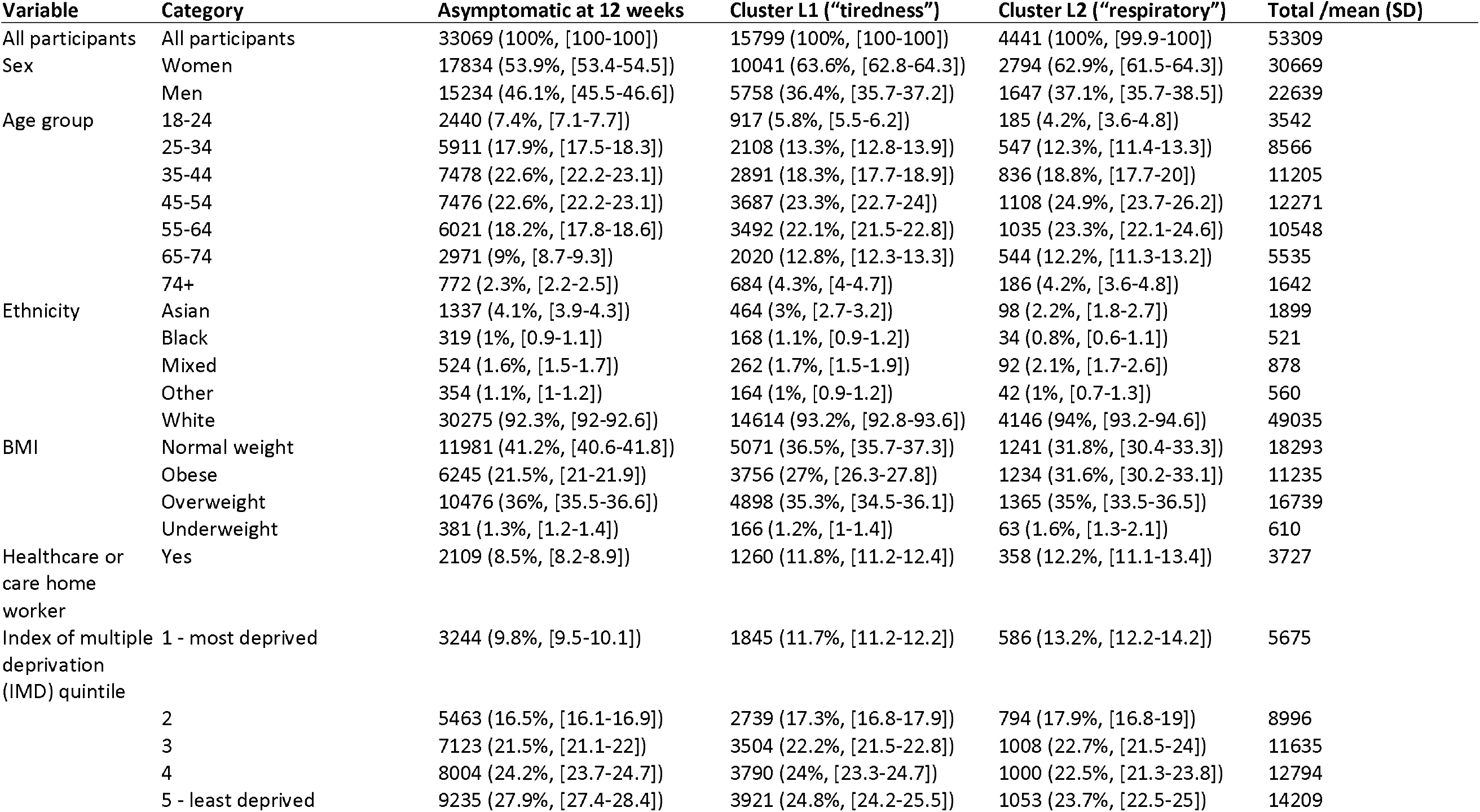
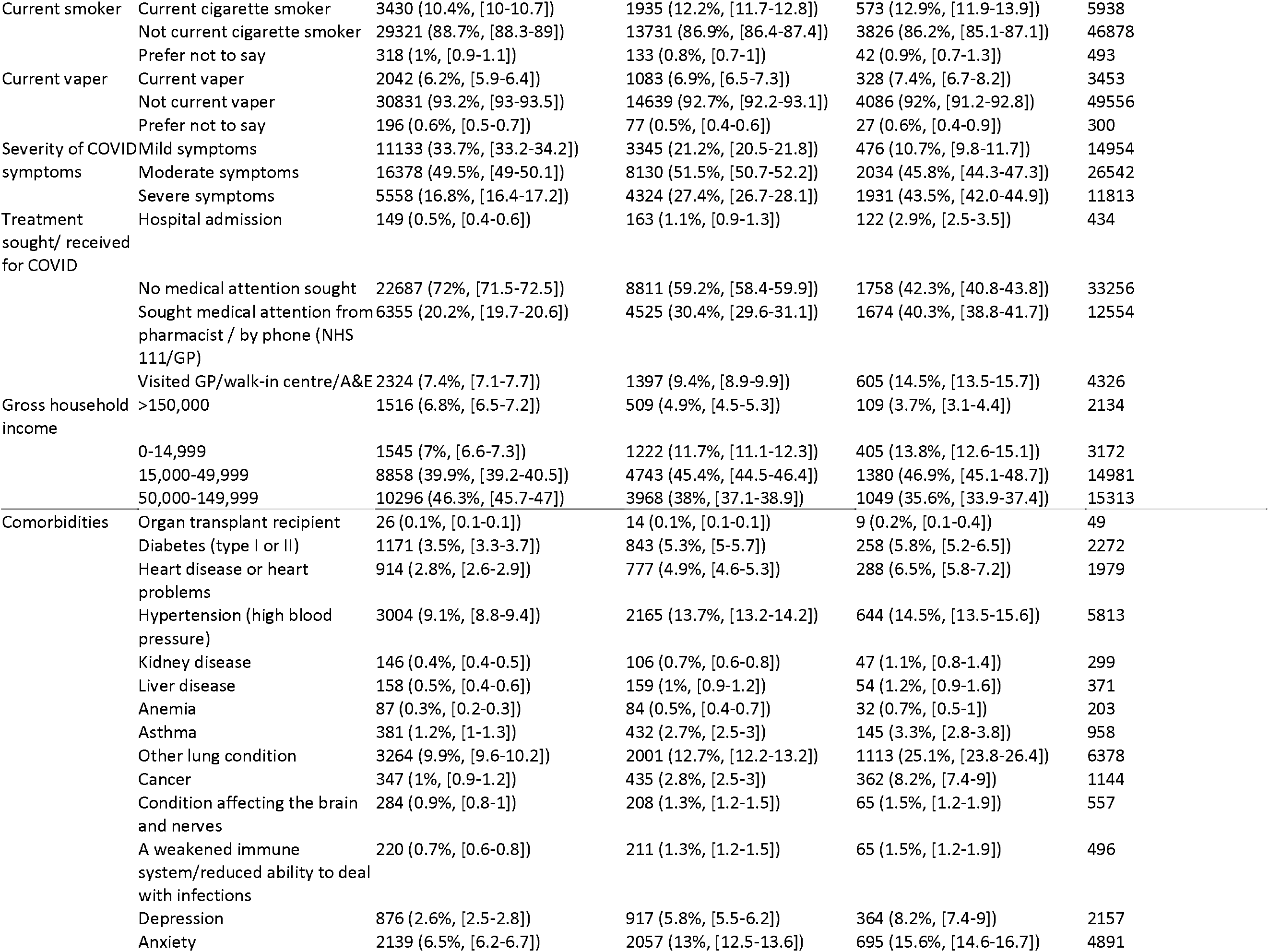
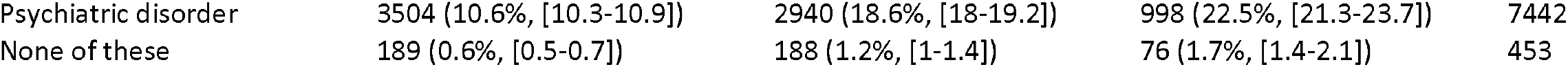
Characteristics of 12-week symptom clusters among 70% sub-sample (n=53,309) of people with symptoms and date of symptom onset 12 weeks or more before survey date --compared with those who were not experiencing any persistent symptoms at 12 weeks. For categorical variables, column-wise within-group percentages shown in parentheses, with 95% confidence intervals in square brackets. Cluster L2 (respiratory) contained proportionately more people who were obese (31.6%, vs 27.0% in Cluster L1), more smokers (12.9% vs 12.2%) and more people who rated their symptoms as ‘severe’ (43.5% vs 27.4%). Prevalence of all surveyed comorbidities was higher in Cluster L2 than Cluster L1, and the prevalence of comorbidities was higher in the symptomatic clusters than in those who did not experience persistent symptoms at 12 weeks.

**Figure S1.**
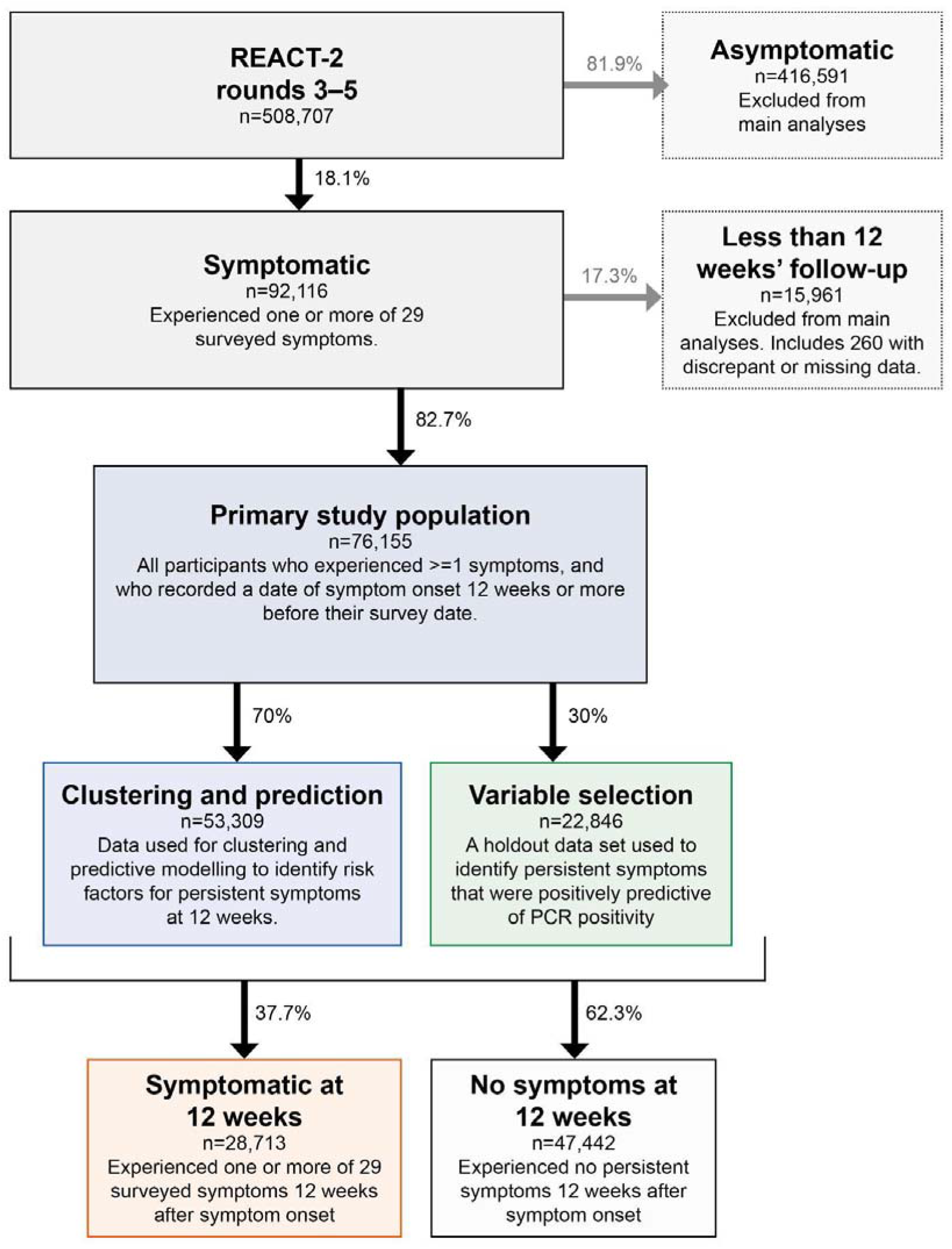
Study population flow chart

**Figure S2.**
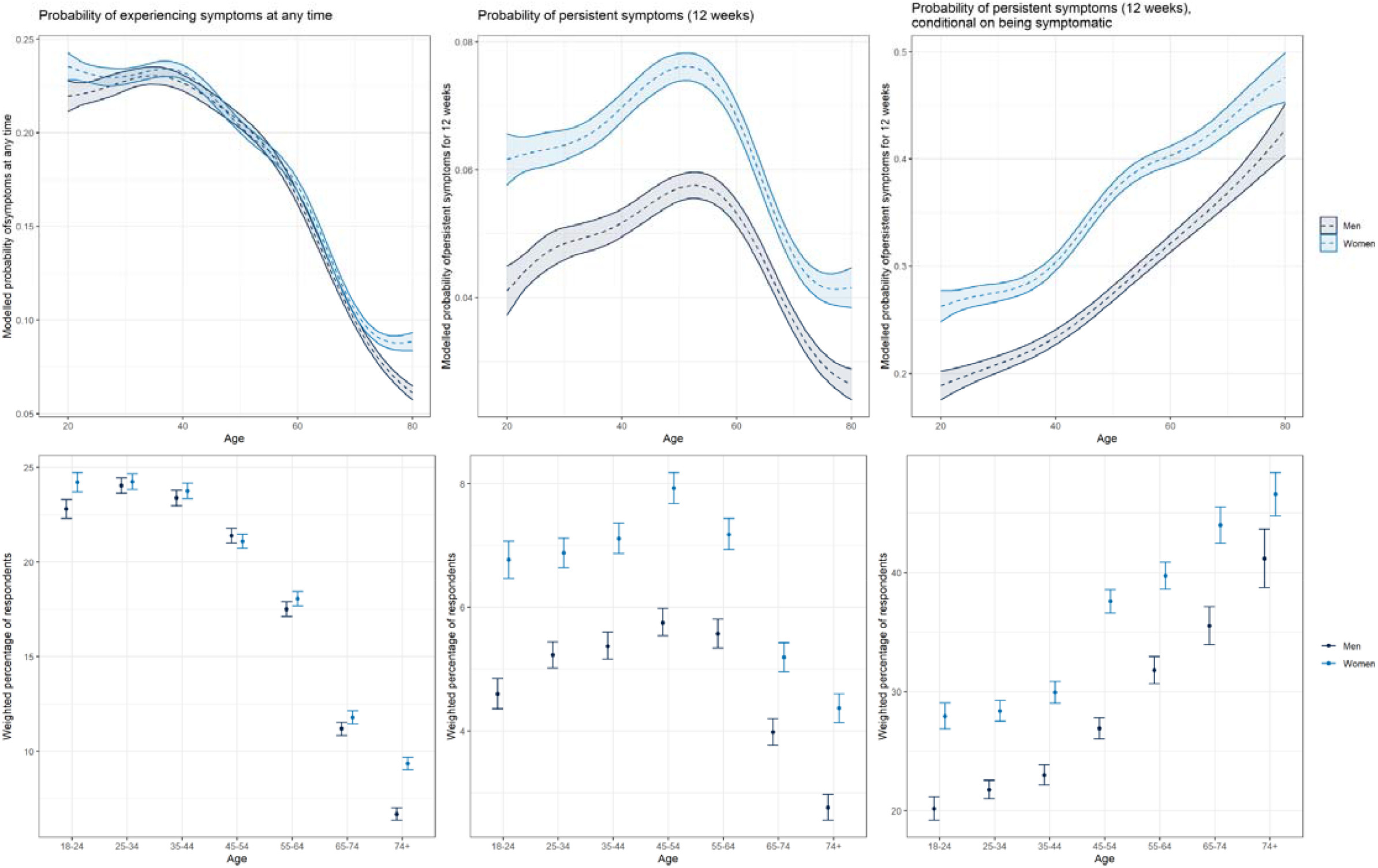
Plots showing (top row) modelled probability of persistent symptoms at 12 weeks as a function of age and sex, using generalised additive models with splines on age and interactions between age and sex (top row), and (bottom row) weighted prevalence of persistent symptoms at 12 weeks. From left to right, plots show (i) probability of experiencing COVID-19 symptoms in the population, (ii) probability of experiencing persistent symptoms at 12 weeks in the population, and (iii) probability of experiencing persistent symptoms at 12 weeks, conditional on symptomatic infection. Conditional on symptomatic infection (likelihood of which decreases after age 50, see central panels), risk of persistent symptoms increases linearly with age (right panels).

**Figure S3.**
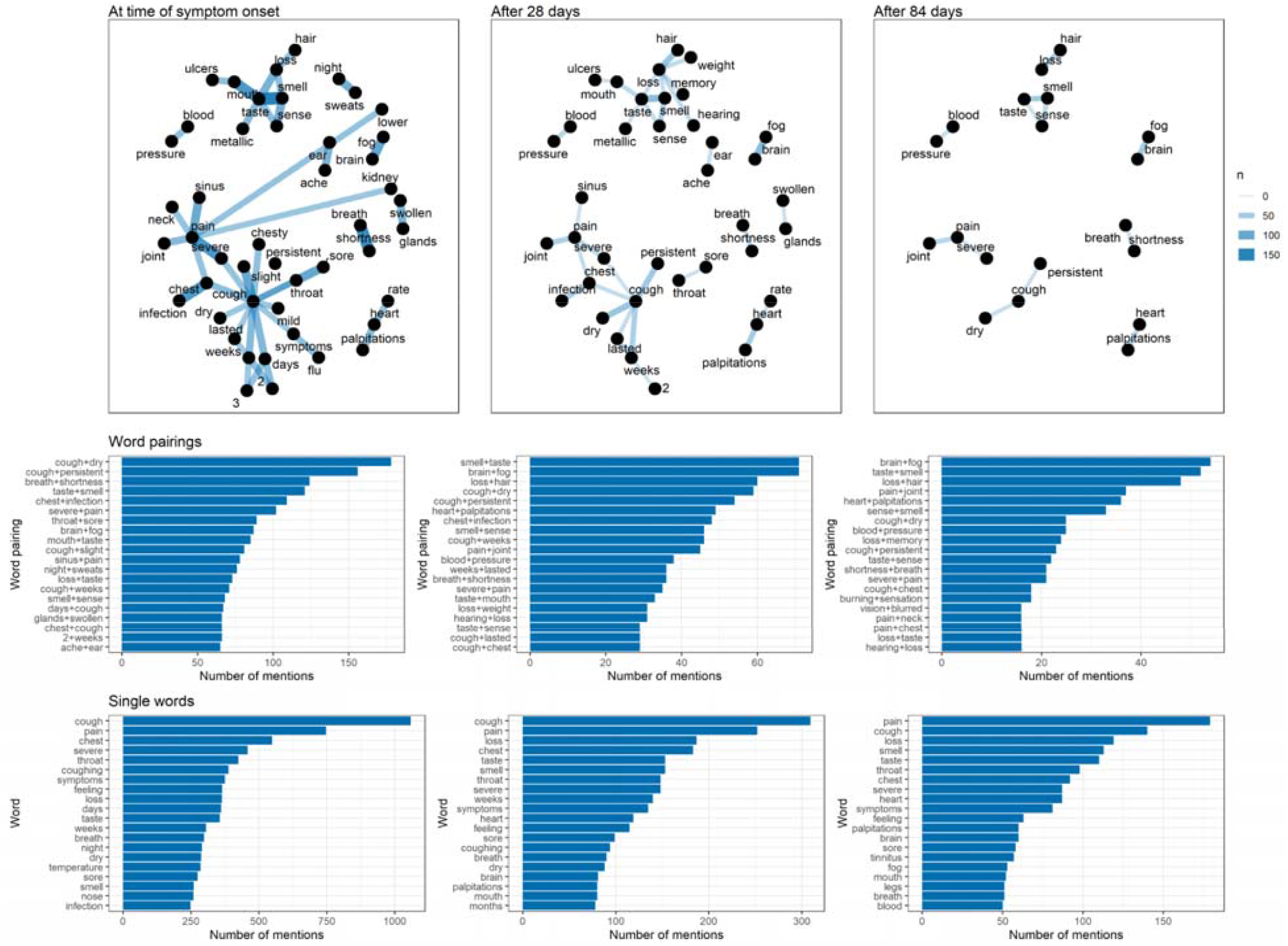
Analysis of the free text responses to COVID-19 symptom questions in REACT-2. A total of 8,374 respondents left free-text responses, of whom 1,860 reported the described symptoms persisting for 12 weeks or more. From left to right, the panels show (i) analysis of symptoms occurring at time of symptom onset; (ii) analysis of symptoms occurring at 4 weeks after symptom onset, and(iii) analysis of symptoms occurring at 12 weeks after symptom onset. The top two rows of panels visualise within-response word co-occurrence, as networks (top panels) and simple bar plots of the top 20 most frequently co-occurring pairs of words (middle panels). The bottom row shows the most commonly occurring single words.

**Figure S4.**
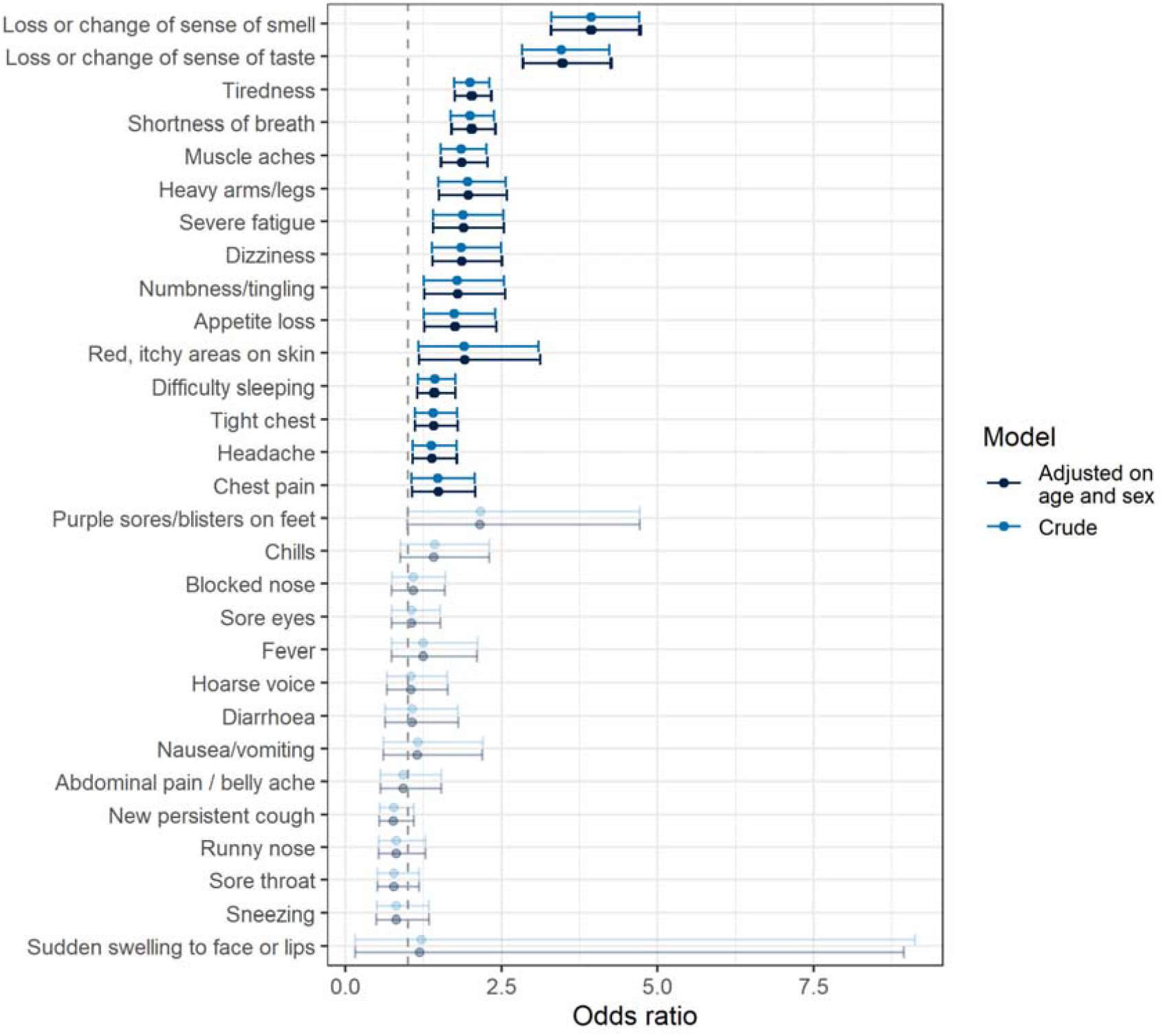
Results of logistic model with self-reported PCR positivity as the binary outcome variable, and the 29 surveyed symptoms as binary independent variables, among a 30% variable selection holdout data set. The 15 variables that were identified as positively predictive of PCR positivity (shown in bold colours) were taken forward as a sensitivity analysis of prevalence of persistent symptoms beyond 12 weeks, and as outcome variables in logistic regression of risk factors for persistence, among this reduced symptom set.

**Figure S5.**
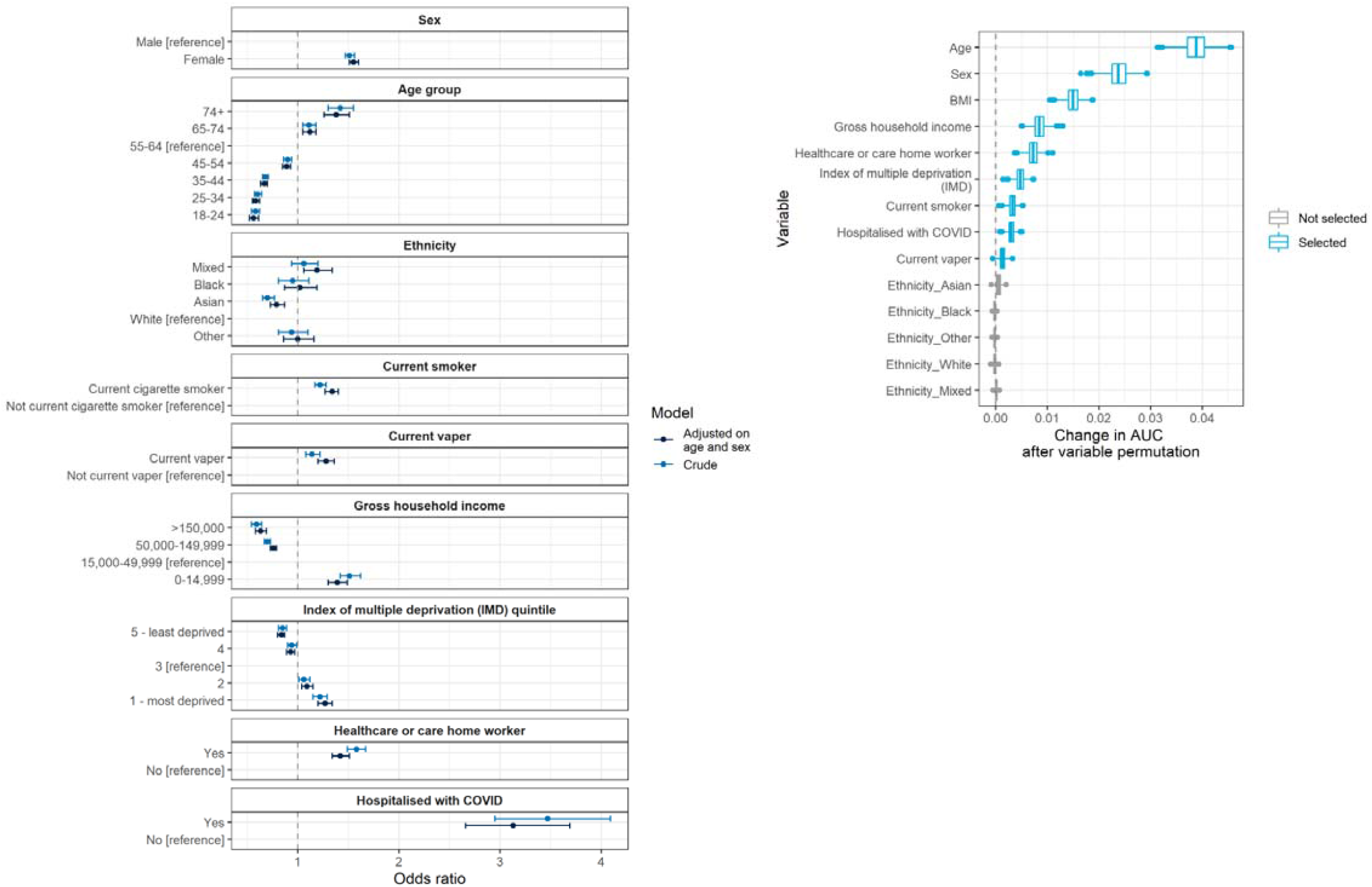
Plots showing the results of predictive modelling, with one or more of a subset of 15 symptoms at 12 weeks (y/n) as the binary outcome variable. Left panel shows odds ratios from logistic regression models, both crude and adjusted on sex and age. Right panel shows the mean contribution in area under the curve (AUC) that each variable makes to a multivariable boosted tree model, derived using variable permutation. Age, sex, BMI, smoking status and prior hospitalisation with COVID-19 are the strongest predictors of persistent symptoms in multivariable modelling, while Asian ethnicity is associated with a lower risk of persistent symptoms at 12 weeks; Asian ethnicity is selected as a predictor in multivariable boosted tree modelling.

**Figure S6.**
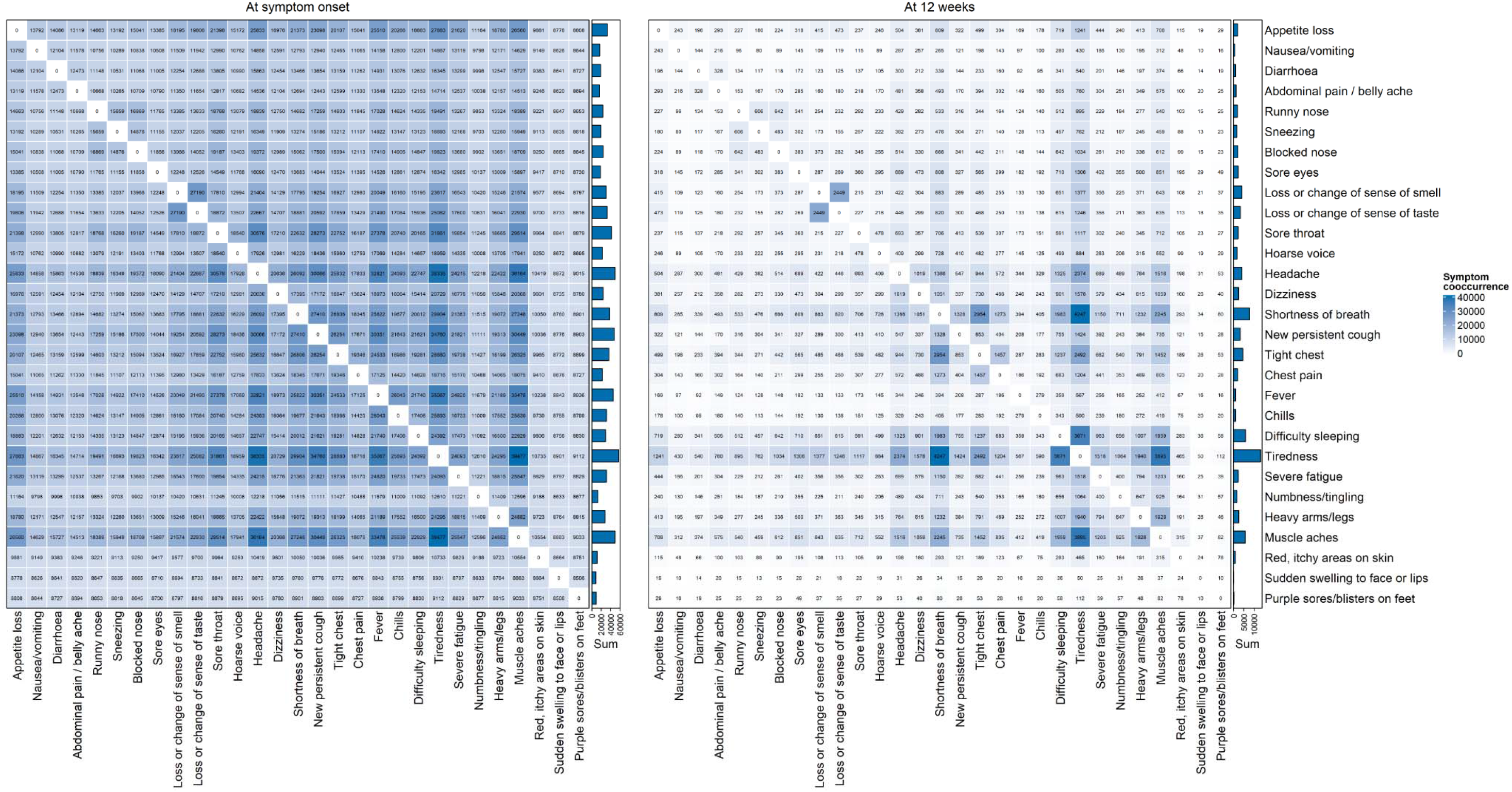
Pairwise co-occurrence heatmaps of symptoms at time of symptom onset (left) and 12 weeks after symptom onset (right), among 76,155 symptomatic participants. Deeper blue colour indicates that more people had both of the paired symptoms at symptom onset or 12 weeks post symptom onset; bar plots indicate marginal counts of participants with each symptom at symptom onset and at 12 weeks post symptom onset.

## Notes

### Competing Interest Statement

The authors have declared no competing interest.

### Clinical Protocols

https://www.imperial.ac.uk/medicine/research-and-impact/groups/react-study/

